# Integration of proteomics with genomics and transcriptomics increases the diagnostic rate of Mendelian disorders

**DOI:** 10.1101/2021.03.09.21253187

**Authors:** Robert Kopajtich, Dmitrii Smirnov, Sarah L. Stenton, Stefan Loipfinger, Chen Meng, Ines F. Scheller, Peter Freisinger, Robert Baski, Riccardo Berutti, Jürgen Behr, Martina Bucher, Felix Distelmaier, Elisabeth Graf, Mirjana Gusic, Maja Hempel, Lea Kulterer, Johannes Mayr, Thomas Meitinger, Christian Mertes, Metodi D. Metodiev, Agnieszka Nadel, Alessia Nasca, Akira Ohtake, Yasushi Okazaki, Rikke Olsen, Dorota Piekutowska-Abramczuk, Agnès Rötig, René Santer, Detlev Schindler, Abdelhamid Slama, Christian Staufner, Tim Strom, Patrick Verloo, Jürgen-Christoph von Kleist-Retzow, Saskia B. Wortmann, Vicente A. Yépez, Costanza Lamperti, Daniele Ghezzi, Kei Murayama, Christina Ludwig, Julien Gagneur, Holger Prokisch

## Abstract

By lack of functional evidence, genome-based diagnostic rates cap at approximately 50% across diverse Mendelian diseases. Here, we demonstrate the effectiveness of combining genomics, transcriptomics, and, for the first time, proteomics and phenotypic descriptors, in a systematic diagnostic approach to discover the genetic cause of mitochondrial diseases. On fibroblast cell lines from 145 individuals, tandem mass tag labelled proteomics detected approximately 8,000 proteins per sample and covered over 50% of all Mendelian disease-associated genes. Aberrant protein expression analysis allowed the validation of candidate protein-destabilising variants, in addition to providing independent complementary functional evidence to variants leading to aberrant RNA expression. Overall, our integrative computational workflow led to genetic resolution for 22% of 121 genetically unsolved whole exome or whole genome negative cases and to the discovery of two novel disease genes. With increasing democratization of high-throughput omics assays, our approach and code provide a blueprint for implementing multi-omics based Mendelian disease diagnostics in routine clinical practice.

---

Whole exome sequencing (WES) and whole genome sequencing (WGS) resolve the molecular cause of suspected genetic disease in approximately 30-50% of cases (Clark et al., 2018). The overarching challenge of the analysis is the interpretation of the vast number of variants of uncertain significance (VUS). The current ACMG recommendation for interpretation of genetic variants (Richards et al., 2015) attaches high importance to functional validation in designation of a variant as pathogenic (P) or likely pathogenic (LP). For this reason, systematic application of RNA sequencing (RNA-seq) has proven valuable in reducing the diagnostic shortfall of WES/WGS by providing a functional readout of variant consequence on RNA abundance and form, through the detection of aberrant expression, aberrant splicing, and/or monoallelic expression of heterozygous pathogenic variants. In this way, RNA-seq allows the pathogenic designation of synonymous and non-coding VUS. The approach provides a molecular diagnosis to approximately 10% of unsolved WES/WGS cases with a suspected mitochondrial disease (Kremer et al., 2017) and up to 35% in other disease cohorts (Cummings et al., 2017, Gonorazky et al., 2019, Fresard et al., 2019). Proteomics has the added potential to capture the consequence of protein-destabilising missense variants, a variant class eluding detection by RNA-seq. Missense variants are one of the most frequently reported functional classes of P/LP variants in Mendelian disease according to the ClinVar database (Simple ClinVar, see **online Resources**), accounting for just over 25%. While proteomics has been used to elucidate underlying disease mechanism (Lake et al., 2017, Borna et al., 2019, Stojanovski et al., 2020), to provide cumulative functional evidence to RNA-seq candidates (Kremer et al., 2017), and to resolve a small number of cases with suspected monogenic disease by guiding targeted genetic testing (Grabowski et al., 2019), the utility of the systematic application and integration of quantitative proteomics with WES/WGS data into a diagnostic pipeline in a large cohort of unsolved cases has yet to be explored. Here, we establish an integrative multiomic workflow for the parallel analysis of WES/WGS, Human Phenotype Ontology (HPO) descriptors, RNA-seq, and quantitative proteomic data to provide comprehensive capture of variant consequence on the gene product(s) with the objective to genetically resolve WES/WGS unsolved mitochondrial disease cases.

Initiated by the GENOMIT project (see **online Resources**), we have analysed over 2,000 clinically suspected mitochondrial disease cases by WES/WGS and gathered corresponding HPO terms for automated phenotype integration (Stenton et al., 2021). Mitochondrial diseases are a prime example of the diagnostic challenge faced in human genetics given their vast clinical and genetic heterogeneity. In-keeping with previous studies (reviewed by Stenton and Prokisch 2020), we reached a diagnosis by WES/WGS analysis for approximately 50% of the cases. Here, selecting 143 of these mitochondrial disease cases (121 unsolved and 22 solved positive controls) plus two healthy controls with available fibroblast cell lines, we performed RNA-seq and quantitative tandem mass tag (TMT) labelled proteomics in order to take an integrative multi-omic approach to diagnosis (**Fig. 1a**, Supplementary Fig. 1a-c) (see **online Methods**).

**Fig. 1:**
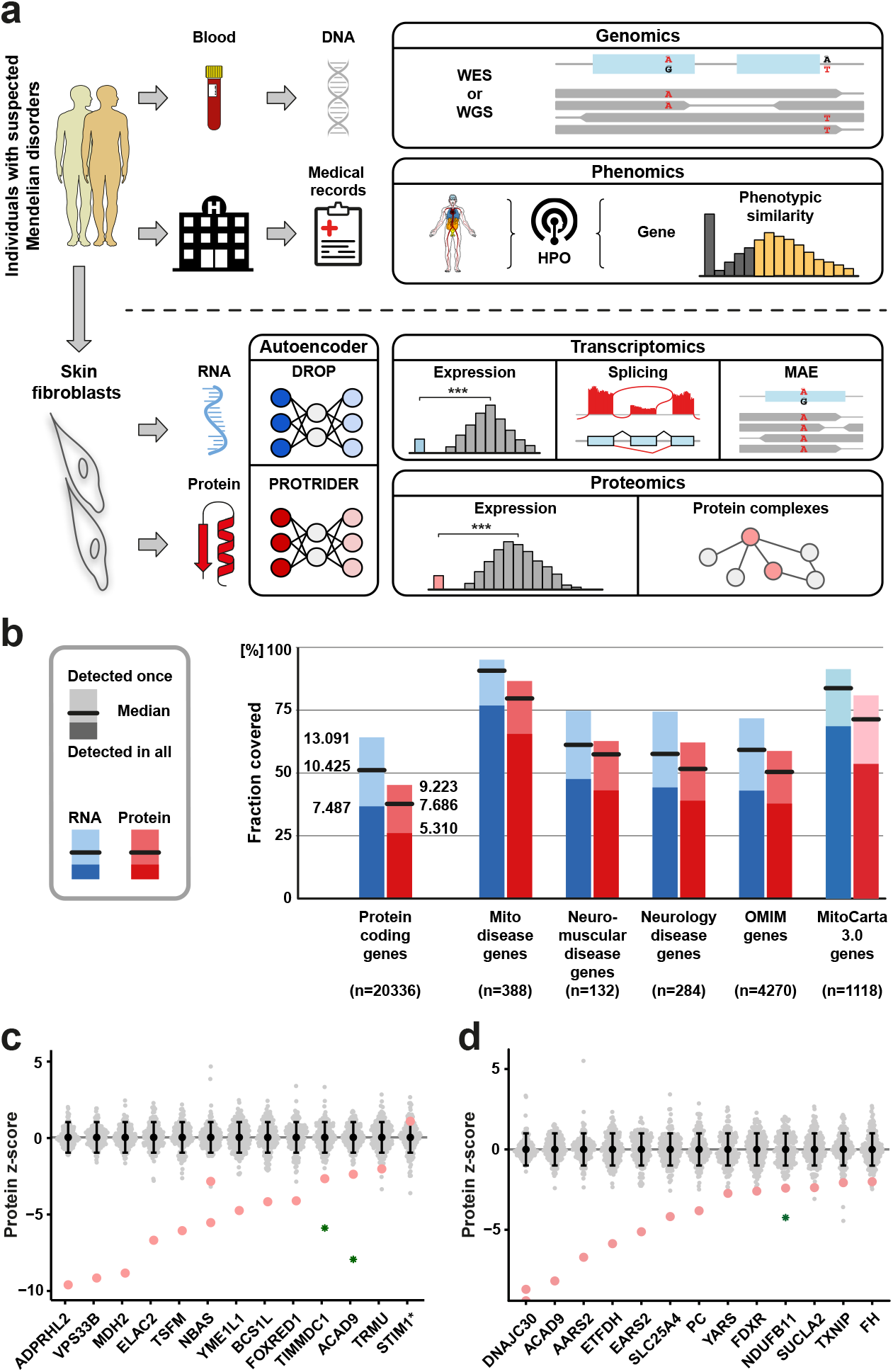
Genetic diagnosis by simultaneous genomic (WES/WGS), phenomic, transcriptomic (RNA-seq), and proteomic investigation followed by integrated analysis. **a**, Multi-omic approach based on the integration of genomics (WES/WGS), transcriptomics (RNA-seq), proteomics, and phenotypic descriptors (HPO). We obtained DNA for WES/WGS from blood and RNA-seq and proteomics from fibroblasts (acquired by minimally invasive skin biopsy). Functional evidence from each omic was integrated in search of a genetic diagnosis. The resultant diagnoses were thereby supported by multiple lines of robust clinical and functional evidence. Simultaneously, heterozygous and potentially biallelic genetic variants were prioritized according to their effect on the corresponding transcript(s) and protein by the identification of aberrant expression in RNA-seq and proteomic data, in addition to aberrant splicing and monoallelic expression (MAE) of a deleterious heterozygous variants in RNA-seq data. Phenotype data complemented the analysis by gene-level prioritization based upon semantic similarity scoring. Together, omics integration allowed comprehensive gene-variant prioritization by providing insight into the effect of rare variation on expression of gene products. An overview of our multiplexed, time-efficient, RNA-seq and proteomic sample workflow is depicted in **Supplementary Fig. 1a-c. b**, Proportion of protein-coding genes detected by RNA-seq (blue) and proteomics (red), genome-wide and among mitochondrial disease genes (Schlieben and Prokisch 2020), Neuromuscular and Neurology genes (Frésard et al., 2019), and OMIM disease genes (https://omim.org), in addition to proteins predicted to localise to the mitochondrial organelle according to MitoCarta 3.0 (Rath et al., 2020). **c**, Protein z-score distribution for disease-causing genes in WES/WGS solved cases (positive controls). **d**, Protein z-score distribution for disease-causing genes with prioritized variants in WES/WGS unsolved cases. In panels **c** and **d**, the points appear in red for validated cases and in green for novel cases diagnosed in our downstream systematic discovery approach.

With the detection of approximately 12,000 transcripts and 8,000 proteins per sample (**Supplementary Fig. 1d**), a median of 91% (n=353) and 80% (n=310) of mitochondrial disease gene products, and 59% (n=2,535) and 51% (n=2,159) of all Mendelian disease gene products were quantified per sample in RNA-seq and proteomics, respectively. These figures deem fibroblasts an easily accessible tissue with high disease gene coverage and an excellent resource for the study of mitochondrial diseases (**Supplementary Fig. 1e**).

To validate our quantitative proteomic approach on fibroblast cell lines, we first asked whether we were able to detect protein underexpression in 18 positive controls with pathogenic nuclear-encoded variants and and four positive controls with pathogenic mtDNA-encoded variants known to lead to protein destabilisation (i.e., cases with previous immunoblotting demonstrating reduced protein level). Our proteomic approach validated 14 cases (14/22, 64%) by nominally significant protein underexpression (**Fig. 1c**). Of the cases unable to be validated (8/22), in three the protein was not detected and in five there was no significant change in protein expression, this included all four mtDNA-encoded variants (**Supplementary Table 1**).

Next, in our cohort of 121 unsolved cases we investigated 21 where clinical analysis of the WES data had resulted in a diagnostic report of a rare VUS in a known OMIM disease gene, according to the ACMG criteria (**Supplementary Table 2**, for case summaries see **Supplementary Information**). These 21 cases carried a total of 26 unique VUS, the majority of which (20/26, 78%) were missense in nature. Variant pathogenicity was validated in 14 of the 21 cases by nominally significant protein underexpression (**Fig. 1d**) and in one case by aberrant RNA splicing (total, 15 cases, 71%). In total, 4/14 of the diagnoses validated by proteomics were also validated by reduced RNA expression levels (**Supplementary Fig. 1f** and **Supplementary Table 2**).

We were unable to provide functional evidence of pathogenicity in six of the 21 cases. The reasons for this are manifold. First, the variant may be pathogenic but exert pathogenicity by a defect in protein function and not in protein stability, a consequence which cannot be captured by protein underexpression. Second, the variant may indeed not be pathogenic, exemplified in individual OM00762, where proteomics was valuable in rejecting the prioritised variant in the mitochondrial targeting sequence of *MRPL53* due to normal expression of both MRPL53 and the large mitoribosomal subunit (**Supplementary Fig. 1g**) and in individual OM36526, where the prioritised homozygous variant in *VPS11* could be rejected and an alternative genetic diagnosis in *MRPS25* discovered. Third, the protein may not be detected in the respective proteomics batch, as exemplified by C19ORF70 in individual OM66072 (**Supplementary Table 2**).

To identify genes with aberrant RNA expression, we performed three outlier analyses, i) aberrant expression levels, ii) aberrant splicing, and iii) monoallelic expression of rare variants via the DROP pipeline (Yépez et al., 2021). To identify aberrant protein expression, we developed the algorithm PROTRIDER which estimates deviations from expected protein intensities while controlling for known and unknown sources of proteome-wide variation (see **online Methods**). We compared PROTRIDER to z-scores calculated after regressing out covariates (instrument, sample preparation batch, TMT batch, and gender) as well as an approach based on testing for differential expression of each sample against all others. As outlier calls from PROTRIDER were more enriched for rare stop, frameshift, splice, and missense variants and PROTRIDER achieved a better fit of the data, we decided to adopt PROTRIDER for aberrant protein expression detection (see **online Methods**).

Our matched genome, transcriptome, and proteome datasets together with protein expression outlier calls allowed us to investigate how aberrant RNA and protein expression relate to one another in the context of rare genetic variation, to our knowledge for the first time. After multiple-testing correction, we identified a median of two aberrantly expressed transcripts and six aberrantly expressed proteins per sample (**Supplementary Fig. 2a**). The majority of RNA outliers resulted in protein outliers (77%) (**Fig. 2a**). Those not resulting in a protein outlier may be explained by buffering mechanisms on the protein level (Ishikawa et al., 2017, Battle et al., 2015, Vogel et al., 2012), artefact, or lack of power.

**Fig. 2:**
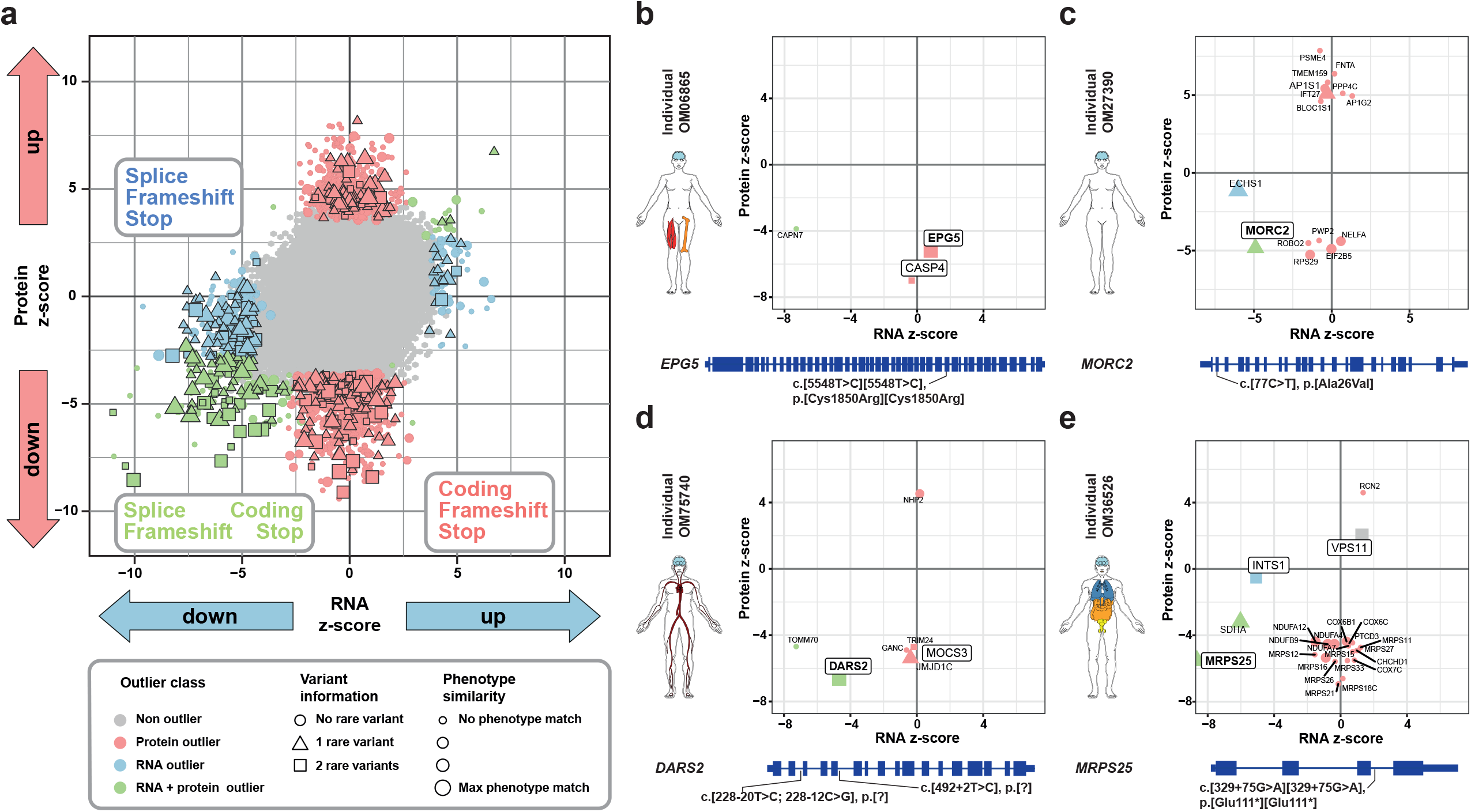
Application of an integrative multi-omic approach to detect the pathogenic consequence of genetic variants. **a**, RNA z-scores (x-axis) vs. protein z-scores (y-axis) detected by RNA-seq and proteomics across all samples. The shape indicates rare variants (minor allele frequency <1%) in the encoding gene. The size indicates semantic similarity with the established disease-gene associated phenotype. The colour represents outlier class. All detected splice defects and MAE events resulted in aberrant expression, allowing this to be used as an indicator of variant pathogenicity. All splice, stop, and frameshift variants in RNA-only expression outliers (n=37), were observed in heterozygous state. **b**, Individual OM06865 presented in childhood with predominantly neurological and muscular involvement. A homozygous missense variant in the autosomal recessive disease gene *EPG5* was prioritised as a protein-only outlier. **c**, Individual OM27390 presented in infancy with failure to thrive, global developmental delay, seizures, encephalopathy, nystagmus, hypotonia, and abnormality of the basal ganglia on MRI. A heterozygous missense variant in the autosomal dominant disease gene *MORC2* was prioritized by an RNA-and-protein outlier. **c**, Individual OM75740 presented in infancy with muscular hypotonia, cardiomyopathy, and abnormalities in the cerebral white matter on MRI. *DARS2* was detected as an RNA-and-protein underexpression outlier explained by a combination of splice and intronic variants. **d**, Individual OM36526 presented in childhood with global developmental delay, elevated lactate, and an isolated respiratory chain complex (RCC) IV defect. The MRPS25 RNA-and-protein outlier is explained by a homozygous intronic variant demonstrating aberrant splicing. In addition, three other proteins from the small mitoribosomal subunit were underexpressed as a downstream consequence of the primary defect.

The expression outliers were stratified into three classes: RNA-only, protein-only, and RNA-and-protein outliers. Though overexpression can provide evidence for impaired function (Ross and Poirier, 2004), we chose to focus on the two thirds of outliers that are underexpressed, as a clear functional consequence of pathogenic variants in Mendelian disease. All three classes of underexpression outliers were significantly enriched for rare variants in their encoding gene (**Supplementary Fig. 2b-d**). Stratifying for variant function, the enriched variants were splice, stop, and frameshift in nature. These findings are in line with other RNA outlier studies (Li et al., 2017, Ferraro et al., 2020). In addition, protein-only outliers captured the functional consequence of missense variants and in-frame indels, thereby demonstrating significant enrichment for coding variants. Overall, substantially more RNA-and-protein outliers (15%) could be explained by potentially biallelic rare variants, as compared to RNA-only and protein-only outliers (approximately 5%, respectively) (**Supplementary Fig. 2c**). An additional 25% of RNA-and-protein outliers were associated with rare heterozygous variants. Protein outliers without rare variants in the encoding gene may be explained indirectly as a consequence of protein complex instability due to a defect in one of the interaction partners, such as in RCCI (Kremer et al., 2017) or the mitoribosomal complex (Lake et al., 2017, Borna et al., 2019), and by downstream consequences, for example, reduced protein biogenesis secondary to a mitoribosomal protein defect (Lake et al., 2017, Borna et al., 2019). Collectively, these genome-wide observations emphasize the complementarity of proteomics to RNA-seq in capturing the functional impact of rare genetic variation. Moreover it shows the sensitivity of the approach not only to biallelic variation, a hallmark of a recessive inheritance mode, but also to mono-allelic variation, i.e., those responsible for dominant diseases. If these biallelic or single variants are in-keeping with the inheritance mode and phenotype of the known disease gene, the detection of an outlier may lead to diagnosis of the patient.

Aiming to pinpoint pathogenic variants for the remaining unsolved cases (n=106), we next combined the aberrant expression analysis with patient HPO annotations (**Fig. 2a, Supplementary Fig. 2d-e**). Focussing on multiple-testing corrected significant RNA and/or protein underexpression outliers (median four per sample), a median of one outlier matched with the patient phenotype as described by HPO annotations (n=155 outliers in total) or carried rare variant(s) in the corresponding gene (n=177 outliers in total) (see **online Methods**).

Manual inspection and interpretation of outliers resulted in the diagnosis of 12 cases (11%) either in accordance with the ACMG criteria or by functional validation of two novel disease genes, upon identification of the causative variants (**Supplementary Table 3, Fig. 2b-e**, for case summaries see **Supplementary Information**). Of these 12 solved cases, eight were solved by the detection of an RNA-and-protein outlier and four were solved by the detection of a protein-only outlier (**Fig. 2b**).

First considering the eight cases solved by RNA-and-protein outlier detection, one case (OM21111) had a one exon deletion in *NFU1* identified by follow-up WGS in compound heterozygosity with a missense variant resulting in 21% residual protein (z-score −7.8). One case (OM27390) has a heterozygous missense variant in *MORC2* resulting in 69% residual protein (z-score −4.5) (**Fig. 2c**). Four cases demonstrated aberrant splicing resulting in protein outliers, included a homozygous near splice variant in *MRPL44* (z-score −5.5) (OM03592), deep intronic variants in *TIMMDC1* (z-score −6.2) (OM38813) and *MRPS25* (z-score −4.8) (OM36526), and in one case of a direct splice variant on one allele and a unique combination of two frequent intronic variants on the second allele (allele frequency 7.2% and 21.8%, respectively) causing exon skipping in *DARS2* (z-score −6.3) (OM75740) (**Fig. 2d**). The MRPS25 case (OM36526) exemplifies the added value of proteomics in detecting the consequence of a defect on the protein complex given underexpression of five subunits of the small mitoribosomal subunit (**Fig. 2e**). In the final two RNA-and-protein outlier cases, our integrated omics approach led to the identification of novel mitochondrial disease genes, *MRPL38* and *LIG3*.

The MRPL38 outlier (RNA z-score −6.05, protein z-score −5.04) (**Fig. 3a**) illuminated a pathogenic 5’UTR deletion (**Fig. 3b**). The functional relevance was confirmed by reduced abundance of the large mitoribosomal subunit (**Fig. 3c**) resulting in a severe reduction in mitochondrial translation rate rescued by the re-expression of wild-type *MRPL38* (**Fig. 3d**). The LIG3 outlier (RNA z-score −3.26, protein z-score −4.50) (**Fig. 3e**) reprioritised a heterozygous nonsense variant within the mitochondrial targeting sequence (**Fig. 3f**) affecting only the mitochondrial isoform *in trans* with a deep intronic variant causing aberrant splicing. As a dual localized nuclear and mitochondrial DNA ligase, a defect in LIG3 was expected to impact mitochondrial DNA replication, supported by mtDNA depletion and a combined OXPHOS defect in the muscle biopsy (**Fig. 3g**), significantly decreased protein levels of mtDNA encoded gene products (**Fig. 3h**), and impaired mtDNA repopulation (**Fig. 3i**). The downstream functional consequence of the *LIG3* variants was reflected by 63 additional protein outliers.

**Fig. 3:**
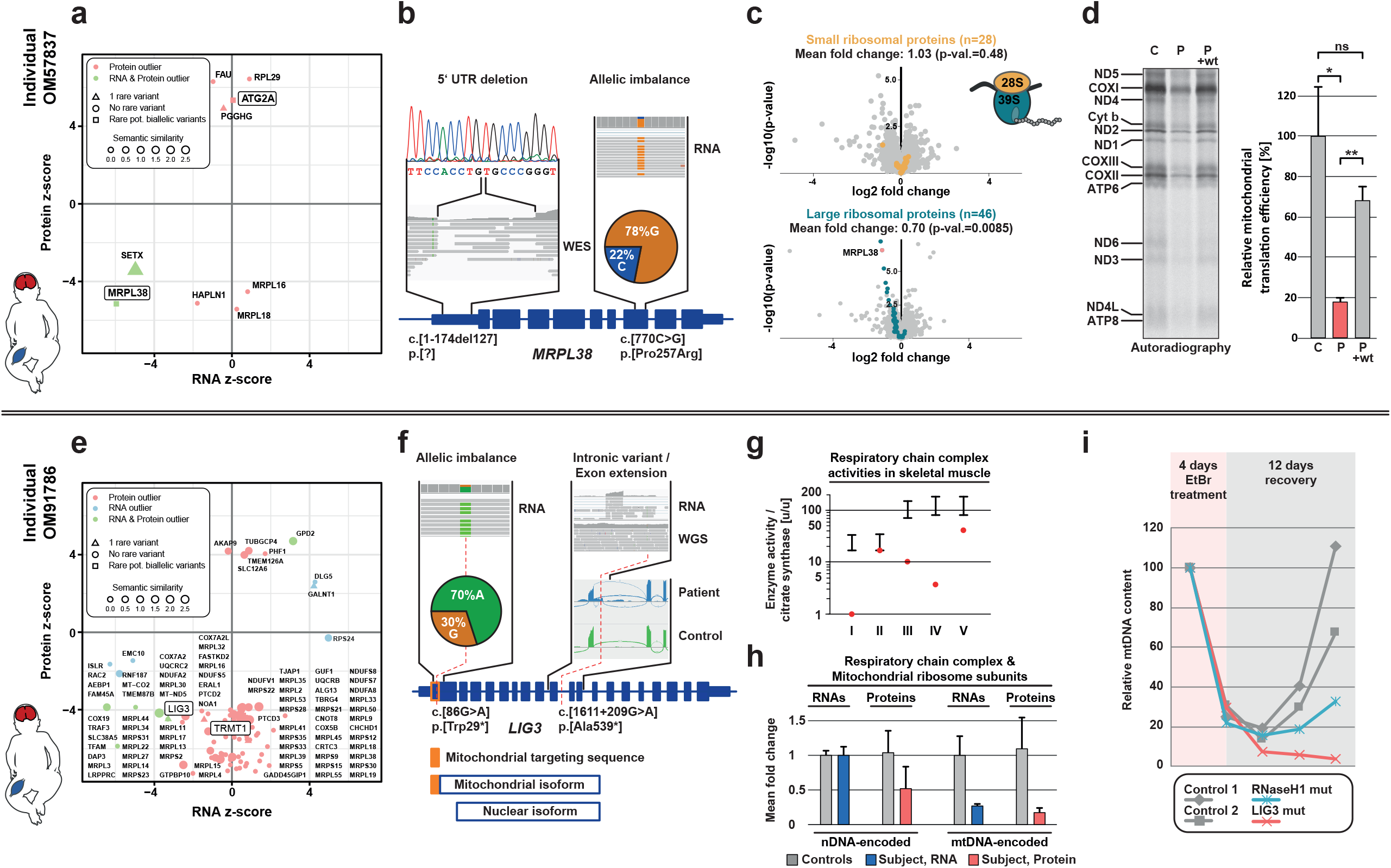
Multi-omic prioritization and functional characterization of two novel mitochondrial disease genes *MRPL38* and *LIG3*. **a**, Individual OM57837 presented in infancy with global developmental delay, intellectual disability, seizures, hypotonia, symmetrical basal ganglia and brainstem abnormalities on brain MRI, and respiratory chain complex (RCC) I and IV defects. The RNA and the protein products of *MRPL38* (Mitochondrial Large Ribosomal Subunit Protein L38) were detected as underexpression outliers. **b**, A missense variant, c.[770C>G], p.[Pro257Arg], present in 78% of RNA reads indicated reduced expression of a compound heterozygous 127 bp deletion in the 5’UTR of *MRPL38*. **c**, Underexpression of MRPL38 resulted in reduction of the large mitoribosomal subunit (n=46 detected subunits). Meanwhile, the small mitoribosomal subunit remained unchanged (n=28 detected subunits). Data are displayed as a gene-wise protein expression volcano plot of nominal (-log_10_) p-values against protein intensity log_2_ fold change. **d**, Measurement of mitochondrial translation in cultured patient-derived fibroblasts by metabolic labelling with [35S]-containing amino acids. The *MRPL38* mutant (P) showed a significantly reduced mitochondrial translation rate compared to control fibroblasts (C). **e**, Individual OM91786 presented with neonatal-onset severe encephalopathy, seizures, hypotonia, and increased serum lactate with early demise in the first weeks of life. LIG3 was identified as an RNA-and-protein outlier. **f**, Whole-genome sequencing identified compound heterozygous variants in *LIG3*. A nonsense variant within the mitochondrial targeting sequence affecting only the mitochondrial isoform of the protein in compound heterozygosity with a deep intronic variant leading to aberrant splicing and partial degradation of the transcript from this allele, indicated by allelic imbalance. **g**, A combined OXPHOS defect sparing nuclear encoded RCCII was present on the muscle biopsy. The black bars represent the reference range. The red point represents the patient measurement. **h**, Normal transcript level but reduced protein level of nuclear-encoded RCC subunits and ribosomal subunits (n=133) indicated complex instability. RNA-and-protein levels of the 13 mtDNA encoded RNAs (11 mRNAs and 2 rRNAs) and 13 mtDNA encoded proteins were reduced. Expression is depicted as a mean-fold change compared to the mean of all other fibroblast samples. **i**, mtDNA copy number in cultured fibroblasts was investigated by qPCR during ethidium bromide induced depletion and repopulation. Impaired mtDNA repopulation was more severe than in the *RNASEH1* mutant cell line which serves as a control for a repopulation defect (Reyes et al. 2015).

Next, looking at the four cases solved by protein-only outliers, in all cases at least one of the responsible variants was missense or inframe indel in nature, variant classes with pathogenic consequence eluding detection by RNA-seq (**Supplementary Table 3**). Amongst these, was a hemizygous X-linked *NDUFB11* missense variant resulting in aberrant protein underexpression (z-score −4.52) and pathologically low abundance of respiratory chain complex (RCC) I (63%) with no rare variants within any other RCCI subunit. The reduction in RCCI was most pronounced in the ND4-module to which NDUFB11 belongs (44% remaining, lowest in dataset), in-keeping with a second confirmed *NDUFB11* case (55% remaining, second lowest in dataset). The detection of this variant in the unaffected grandfather indicated incomplete penetrance. Attributing pathogenicity to variants of incomplete penetrance, even in the presence of a phenotypic match, is an outstanding challenge in human genetics. However, in cases where reduced activity is causative of disease, proteomics has the power to classify variants affecting protein complex abundance. This was also demonstrated for the homozygous variant in *DNAJC30* in two cases, by providing evidence for the loss-of-function character of an incompletely penetrant missense variant (**Supplementary Table 2**), as recently reported in a cohort of 27 families (Stenton et al., 2021).

To summarise, leveraging on advanced proteomics we quantified a substantial fraction of expressed proteins, determined their normal physiological range in fibroblasts, and called protein outliers in a robust manner. By developing an integrated multi-omic analysis pipeline, we establish a clinical decision support tool for the diagnosis of Mendelian disorders. The power of our multi-omic approach is demonstrated by validation and detection of the molecular diagnosis in 27 of 121 (22%) unsolved WES/WGS cases (**Fig. 4a**). Specifically, proteomics had high value in providing functional evidence for protein destabilising missense variants, the most frequent form of P/LP variant in mitochondrial disease and both VUS and P/LP variant reported in ClinVar (**Fig. 4b**), which cannot be interpreted by RNA-seq analysis. Proteomics was thereby essential to the diagnosis of 14 of 27 successfully diagnosed cases (52%). Moreover, in 19 of a total of 49 diagnosed cases in the cohort (38%) we detect downstream functional evidence of variant pathogenicity on the complex level (**Fig. 4a**). Our code is freely available (https://prokischlab.github.io/omicsDiagnostics/) and an interactive web interface allows the user to browse all results and could serve as a basis for developing future integrative multi-omics diagnostic interfaces. We used TMT-labelling, a proteomics technique quantifying the very same peptides for all samples of a batch. This greatly facilitates detection of underexpression outliers compared to conventional untargeted mass-spectrometry which suffers from widespread missing values in low intensity ranges. Though RNA-seq did not allow interpretation of missense variants, it provided independent cumulative evidence and guided the identification of causative splice variants in half of the solved cases. Moreover, RNA-seq has a deeper coverage of expressed genes in comparison to proteomics, capturing 50% more genes. It therefore remains useful for lowly expressed proteins. To identify novel diagnoses we applied stringent significance filtering (FDR<0.1) and focussed on underexpression outliers with a phenotype match or carrying rare variants in the corresponding gene, leading to one protein outlier per sample in median. However, with the integration of multiple levels of omics information and phenotype descriptors, relaxed significance thresholds may in future be considered. Our approach depends on available tissue, encouraging clinicians to be proactive and opportunistic in biosampling (e.g, a minimally invasive skin punch biopsy to establish a fibroblast cell line), specifically when follow-up visits are unlikely. Given the increasing democratization of proteomics we envisage its implementation in clinical practice to advance diagnostics by routine integration of functional data.

**Fig. 4:**
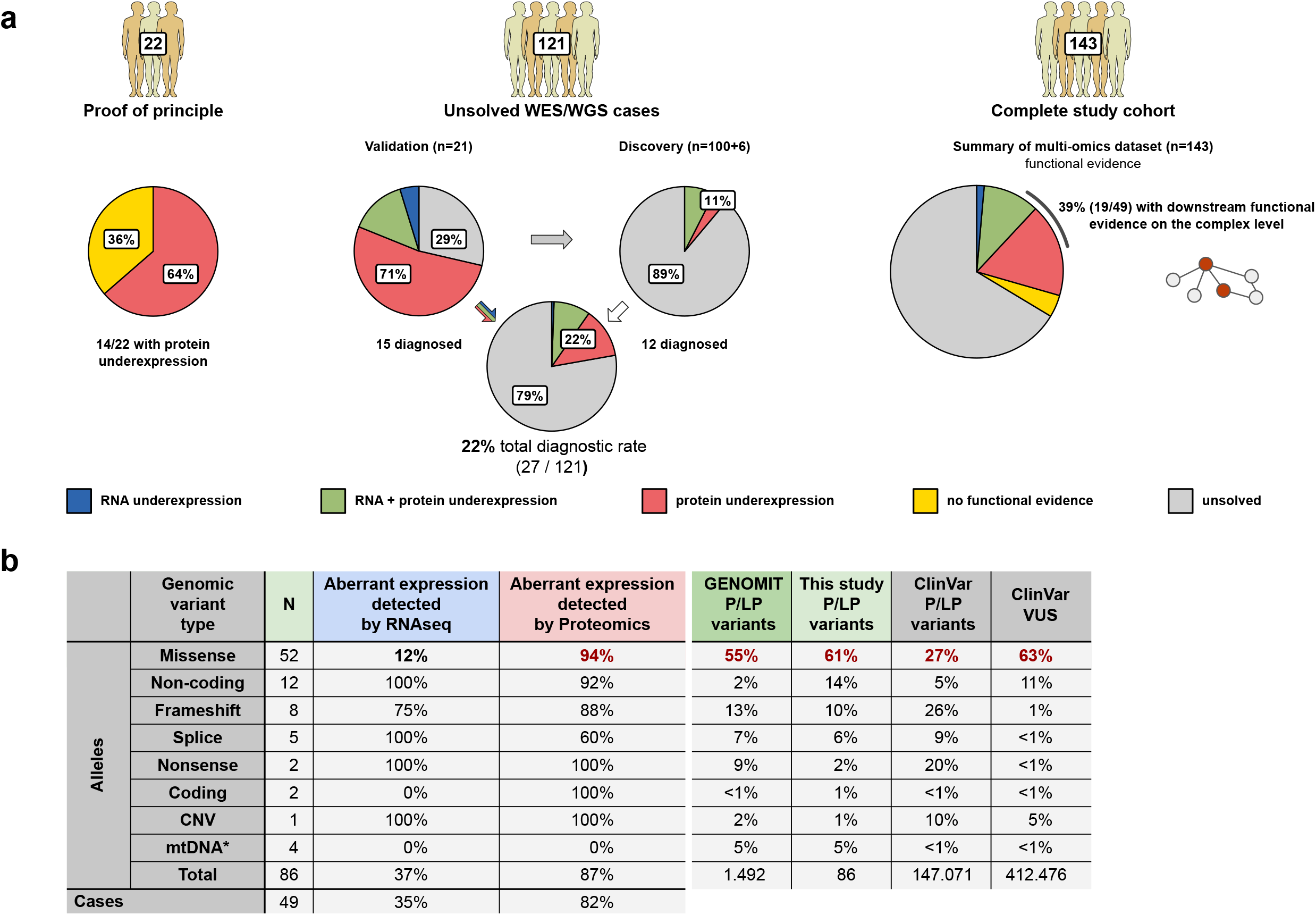
The power of an integrative multi-omic approach to detect the pathogenic consequence of genetic variants. **a**, A summary of the functional evidence for pathogenicity provided by multi-omics in the study cohort. In the proof of principle approach in WES/WGS solved cases (n=22), functional evidence of pathogenicity was provided by protein underexpression in 14/22 patients (64%). In the validation approach, VUS were confirmed to be P/LP in 15/21 unsolved cases (71%). In the discovery approach, 12 novel diagnoses were made in 12/106 unsolved cases (11%). Together, this resulted in an overall diagnostic rate of 22% when applying a multi-omics pipeline to unsolved WES/WGS cases in diagnostics. Overall, of the total of 49 molecularly diagnosed cases in the cohort, proteomics revealed downstream functional evidence of pathogenicity on the complex level in 39% (19/49), accounting for 140/397 underexpression outliers detected amongst these diagnosed cases. **b**, A summary of the variants accounting the patients’ molecular diagnoses in this study with the corresponding functional evidence on the RNA and/or protein level (left) and variants represented within Simple ClinVar, GENOMITexplorer (see **online Resources**) (Stenton et al., 2021), and this study (right), stratified by variant function. Notably, the vast majority of disease-causative variants in our cohort were missense and required proteomics to provide functional evidence of pathogenicity. Given that most ClinVar VUS, ClinVar P/LP, and suspected mitochondrial disease P/LP variants are missense, the importance of proteomics in providing functional evidence of pathogenicity is underlined. *mtDNA variants were missense (n=3) and non-coding (n=1).

## online Methods

### Study cohort

All individuals included in the study or their legal guardians provided written informed consent before evaluation, in agreement with the Declaration of Helsinki and approved by the ethical committees of the centres participating in this study, where biological samples were obtained. This study was completed according to the ethical committee of the Technical University of Munich.

### Cell culture

Primary fibroblast cell lines were cultured as per Kremer et al., 2017. Depending on a material transfer agreement with the referring institution, patient-derived cell lines used in this study can be made available.

### Whole exome sequencing (WES)

Whole exome sequencing was performed as per Kremer et al., 2017 and Zech et al., 2020. SAMtools v.0.1.19 and GATK v.4.0 and called on the targeted exons and regions from the enrichment kit with a +/- 50bp extension.

### Variant annotation and handling

Variant Effect Predictor (McLaren et al., 2016) from Ensembl (Zerbino et al., 2018) was used to annotate genetic variants with minor allele frequencies from the 1000 Genomes Project (1000 Genome Consortium, 2015), gnomAD (Karczewski et al., 2020), and the UK Biobank (Bycroft et al., 2018), location, deleteriousness scores and predicted consequence with the highest impact among all possible transcripts. Variants with minor allele frequency (MAF) less than 1% across all cohorts were considered as rare. Genes harbouring one rare allele were classified as rare, with two or more rare alleles - potentially biallelic. ACMG guidelines for variant classification were implemented for variants in known OMIM disease genes with the InterVar software (Li and Wang, 2015).

### Gene-phenotypic matching

Phenotype similarity was calculated as a symmetric semantic similarity score with the R::PCAN package (Godard and Page, 2016). We considered genes to match phenotypically if the symmetric semantic similarity between the gene and the case HPO annotations was ≥2 (Köhler et al., 2009; Frésard et al., 2019) (**Supplementary Fig. 2d**). Affected organ systems were visualized with R::gganatogram (Maag 2018), based on the patients’ HPO phenotypes corresponding to the third level of HPO ontology (Köhler et al., 2019).

### RNA sequencing

Non-strand specific RNA-seq was performed as per Kremer et al., 2017. Strand-specific RNA-seq was performed according to the TruSeq Stranded mRNA Sample Prep LS Protocol (Illumina, San Diego, CA, USA). Processing of RNA sequencing files was performed as per Kremer et al., 2017.

### Detection of aberrant RNA expression, aberrant splicing, and mono-allelic expression

RNA-seq analysis was performed using DROP (Yepez et al., 2021), an integrative workflow that integrates quality controls, expression outlier calling with OUTRIDER (Brechtmann et al., 2018), splicing outlier calling with FRASER (Mertes et al., 2020), and mono-allelic expression (MAE) with a negative binomial test (Kremer et al., 2017). We used as reference genome the GRCh37 primary assembly, release 29, of the GENCODE project (Frankish et al., 2019) which contains 60,829 genes. RNA expression outliers were defined as those with a false-discovery rate ≤ 0.1. Splicing outliers were defined as those with a gene-level false-discovery rate ≤ 0.1 and a deviation of the observed percent-spliced-in or splicing efficiency from their expected value larger than 0.3. MAE was assessed only for heterozygous single nucleotide variants reported by WES analysis. We retained MAE calls at a false discovery rate ≤ 0.05. Aberrant events of all three types were further inspected using the Integrative Genome Viewer (Robinson et al., 2011).

### Mass spectrometric sample preparation

Proteomics was performed at the BayBioMS core facility at the Technical University Munich, Freising, Germany. Fibroblast cell pellets containing 0.5 million cells were lysed under denaturing conditions in urea containing buffer and quantified using BCA Protein Assay Kit (Thermo Scientific). 15 µg of protein extract were further reduced, alkylated and the tryptic digest was performed using Trypsin Gold (Promega). Digests were acidified, desalted and TMT-labeling was performed according to (Zecha et al., 2019) using TMT 10-plex labelling reagent (Thermo Fisher Scientific). Each TMT-batch consisted of 8 patient samples and 2 reference samples common to all batches to allow for data normalization between batches. Each TMT 10-plex peptide mix was fractionated using trimodal mixed-mode chromatography as described (Yu et al., 2017). LC-MS measurements were conducted on a Fusion Lumos Tribrid mass spectrometer (Thermo Fisher Scientific) which was operated in data-dependent acquisition mode and multi-notch MS3 mode. Peptide identification was performed using MaxQuant version 1.6.3.4 (Tyanova et al., 2016) and protein groups obtained. Missing values were imputed with the minimal value across the dataset.

### Transcriptome-proteome matching

In order to determine the correct assignment of proteome and transcriptome assay from the same sample, we correlated the gene counts with the protein intensities (**Supplementary Fig. 3**). The spearman ranked correlation test was applied to all transcriptome-proteome combinations, using the cor.test function from R. The distribution of the correlation values are plotted and in the case of mismatch two distinctive populations will appear. Correlations >0.2 correspond to matching samples. Only protein intensities >10,000 and genes with ≥50 counts were considered. Protein intensities were log-transformed and centered. RNA counts were normalized by sequencing depth using size factors (Love et al., 2014), log-transformed and centered. The 2,000 genes with the highest dispersion (as computed by OUTRIDER) were selected.

### Detection of aberrant protein expression with PROTRIDER

To detect protein expression outliers while controlling for known and unknown sources of proteome-wide variations, we employed a denoising autoencoder based method, analogous to methods for calling RNA expression outliers (Brechtmann et al., 2018) and splicing outliers (Mertes et al., 2020). Specifically, sizefactor normalized and log_2_-transformed protein intensities were centred protein-wise and used as input to a denoising autoencoder model with three layers (encoder, hidden space, and decoder). All input values were slightly corrupted by adding a small noise term drawn from a Gaussian with mean zero and standard deviation equal to half the standard deviation for each protein. As protein intensities varied strongly between batches, we included the batch as a covariate in the input of the encoder and in the input of the decoder (**Supplementary Fig. 4a-b**). For a given encoding dimension q, we fit the encoder and decoder weights by minimizing the mean absolute error loss to the uncorrupted values over all available non-missing data. The optimal encoding dimension of the autoencoder was determined by artificially injecting outliers and selecting the encoding dimension that yielded the best area under the precision-recall curve (AUPRC) of recovering these injected outliers. For this dimension fitting procedure, artificial outliers were generated with a frequency of 1 per 1000. An outlier log-transformed intensity x°_i,j_ for a sample *i* and a protein *j* was generated by shifting the observed log-transformed intensity x_i,j_ by z_i,j_ times the standard deviation σ_j_ of x_i,j_, with the absolute value of z_i,j_ being drawn from a log-normal distribution with the mean of the logarithm equal to 3 and the standard deviation of the logarithm equal to 1.6, and with the sign of z_i,j_ either up or down, drawn uniformly:

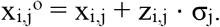

An encoding dimension of 25 was found to be optimal according to this procedure.

After the autoencoder model was fit to the data, statistical testing of the observed log_2_-transformed intensities x_i,j_ with respect to the expected log_2_-transformed intensities μ_i,j_ modelled by the autoencoder was performed, using two-sided Gaussian p-values p_*i,j*_ for sample *i* and protein *j* defined as

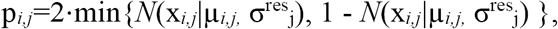

where σ^res^_j_ is the protein-wise standard deviation of the autoencoder residuals x_*i,j*_ - µ_*i,j*_. Finally, p-values were corrected for multiple testing per sample with the method of Benjamini and Yekutieli (Benjamini and Yekutieli, 2001). During the entire process of fitting the autoencoder model as well as the statistical tests, missing data was masked as unavailable and ignored. We refer to this method as PROTRIDER in the following.

### Benchmark of PROTRIDER against alternative approaches

As no method for outlier detection in proteomics data was established yet, we benchmarked our method against an approach that is based on limma (Smyth 2005), which was developed for differential expression analyses on microarray data and assesses statistical significance with a moderated t-statistic. We used recalibrated protein data which has been adjusted with respect to the two identical control samples in each MS-run as the input for limma and included the sex, batch and instrument annotation to adjust for confounding factors. To be able to use limma for outlier detection, we tested each sample against all other samples. Additionally, we compared PROTRIDER to regression of covariates (instrument, sample preparation batch, TMT batch, and gender) followed by z-score calculation on the residuals. We evaluated the performance of all methods based on the proportion of called underexpression outliers that contain a rare stop, frameshift, splice, or missense variant in the gene body of the outlier gene (**Supplementary Fig. 4c**) and overall quality of fit (**Supplementary Fig. 4d-f**). In addition, we compared the ability to recall aberrant protein expression in 28 samples with confirmed pathogenic variants (14 positive controls with pathogenic nuclear-encoded variants and the 14 cases with a validated rare VUS) (**Supplementary Fig. 4c-f**). As PROTRIDER performed favorably in these benchmarks and outlier calls were more enriched for rare variants, we decided to adopt PROTRIDER for the detection of aberrant protein expression.

### Enrichment of genetic variants in outlier genes

We focused our analysis only on the genes where both RNA-and-protein levels were quantified sample-wise and limited it to the genes that were detected as outliers at least once in our cohort. Variants were stratified into six classes according to their impact on the protein sequence, defined by a combination of Ensembl VEP (McLaren et al., 2016) annotations as follows: Stop (stop_lost, stop_gained), splice (splice_region_variant, splice_acceptor_variant, splice_donor_variant), frameshift (frameshift_variant), coding (missense_variant, protein_altering_variant, inframe_insertion, inframe_deletion), synonymous (synonymous_variant, stop_retained_variant), and non-coding (3_prime_UTR_variant, 5_prime_UTR_variant, downstream_gene_variant, upstream_gene_variant, intron_variant, non_coding_transcript_exon_variant, mature_miRNA_variant, intron_variant, intergenic_variant, regulatory_region_variant). Enrichment analysis was performed similarly to described by Li et al 2017, by modelling with logistic regression of each outlier category (RNA only, protein only, and RNA-and-protein over- or underexpresssion) as a function of standardized variant class. For each gene, detected as an outlier of a particular category, the remaining set of individuals served as controls. Proportions of outlier genes were calculated by assignment of one variant class (out of six) with the highest significant enrichment in the corresponding outlier category.

### Detection of aberrantly expressed protein complexes

Detection of aberrantly expressed protein complexes was performed similar to the differential protein complex expression method described by Zhou et al., 2019. Specifically, the quantified proteins were mapped to the protein complex database CORUM (v3.0) (Giurgiu et al., 2019) or to the mitochondria-related subset of HGNC gene groups by gene names. We considered the protein complexes of four subunits or more and with at least 50% of the subunits quantified. For each sample *i* and protein complex *k*, we computed y_i,k_, the mean deviation of observed versus expected protein intensities across all detected subunits (expressed in log_2_ fold-change and as estimated by PROTRIDER or LIMMA). For each protein complex *k*, we fitted by maximum likelihood a Gaussian on all y_i,k_ with mean µ_*k*,_ and standard deviation _*k*_ using the fitdistr function from the R package MASS (Venables and Ripley 2002). The two-sided Gaussian p-values for sample *i* and protein complex *j* was then computed as:

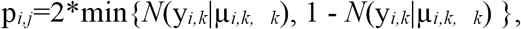

To correct the p-values for multiple testing, the method of Benjamini and Yekutieli (Benjamini and Yekutieli, 2001) was applied per every sample.

### Mitochondrial translation assays

Metabolic labelling of mitochondrial proteins was performed essentially as described previously (Ruzzenente et al., 2018). In brief, patient-derived fibroblasts were incubated in methionine- and cysteine-free DMEM medium supplemented with 10% dialyzed FBS, GlutaMAX, sodium pyruvate (ThermoFisher Scientific, Montigny-le-Bretonneux, France), 100 mg/ml emetine dihydrochloride to block cytosolic protein synthesis and 400 *μ*Ci EasyTag EXPRESS35S Protein Labelling Mix (PerkinElmer, Villebon-sur-Yvette, France). Labelling was performed for 30 min followed by a further incubation for 10 min in standard growth medium. Equal amounts of total cell lysates were fractionated by SDS-PAGE and newly synthesized proteins were quantified by autoradiography.

## Data Availability

The proteomic raw data and MaxQuant search files have been deposited to the ProteomeXchange Consortium (http://proteomecentral.proteomexchange.org) via the PRIDE partner repository and can be accessed using the dataset identifier PXD022803. Code to reproduce the analysis is available via GitHub at github.com/prokischlab/omicsDiagnostics/.

## Data and code availability

The proteomic raw data and MaxQuant search files have been deposited to the ProteomeXchange Consortium (http://proteomecentral.proteomexchange.org) via the PRIDE partner repository and can be accessed using the dataset identifier PXD022803. Code to reproduce the analysis is available via GitHub at github.com/prokischlab/omicsDiagnostics/. All data needed to reproduce the analysis including RNA count and split-read tables as well as the protein intensity matrix are available via Zenodo (Smirnov et al., 2021). The raw sequencing data are not publicly available and no further data is available upon request.

## online Resources

Code to reproduce the figures: https://github.com/prokischlab/omicsDiagnostics/tree/master

PROTRIDER: https://github.com/gagneurlab/OUTRIDER/tree/outrider2

Web interfaces: https://prokischlab.github.io/omicsDiagnostics/#readme.html

Zenodo: https://zenodo.org/record/4501904

PRIDE: https://www.ebi.ac.uk/pride/archive/projects/PXD022803

DROP: https://github.com/gagneurlab/drop

GTEx Portal: https://www.gtexportal.org/home/

OMIM database: www.omim.org

CORUM: https://mips.helmholtz-muenchen.de/corum/

HGNC: https://www.genenames.org

Simple ClinVar: http://simple-clinvar.broadinstitute.org

GENOMIT: https://genomit.eu

GENOMITexplorer: https://prokischlab.github.io/GENOMITexplorer/#readme.html

## Acknowledgements

This study was supported by a German Federal Ministry of Education and Research (BMBF, Bonn, Germany) grant to the German Network for Mitochondrial Disorders (mitoNET, 01GM1906D), the German BMBF and Horizon2020 through the E-Rare project GENOMIT (01GM1920A), the ERA PerMed project PerMiM (01KU2016A), the German BMBF through the e:Med Networking fonds AbCD-Net (FKZ 01ZX1706A), the German Research Foundation/Deutsche Forschungsgemeinschaft (DI 1731/2-2), the AFM-Telethon grant (#19876), the Practical Research Project for Rare/Intractable Diseases from the Japan Agency for Medical Research and Development, AMED (JP19ek0109273, JP20ek0109468, JP20kk0305015, JP20ek0109485), a CMHI grant (S145/16), a PMU-FFF grant (A-20/01/040-WOS), the Pierfranco and Luisa Mariani Foundation (CM23), and the Italian Ministry of Health (GR-2016-02361494, GR-2016-02361241). We would like to thank Caterina Terrile, Franziska Hackbarth, and Hermine Kienberger for their excellent laboratory assistance as well as Miriam Abele for mass spectrometric support at the BayBioMS. We thank the “Cell line and DNA Bank of Genetic Movement Disorders and Mitochondrial Diseases” of the Telethon Network of Genetic Biobanks (grant GTB12001J) and Eurobiobank Network which supplied biological specimens.

## Author Contributions

Conceived and supervised the study, H.P; supervised PROTRIDER development, J.G; performed experiments, R.K, L.K, C.Lu, D.G, Al.N and M.M; analyzed and interpreted results, D.S, S.L, I.S, C.M, V.Y, D.G, M.M, R.K, S.L.S, J.G, H.P; provided essential materials, all authors; wrote the manuscript, S.L.S, H.P, J.G, R.K, and D.S; edited manuscript, all authors.

