## Supplementary Information for "Integration of proteomics with genomics and transcriptomics increases the diagnostic rate of Mendelian disorders"

The Supplementary information contains:

- Supplementary Figures and Legends
- Case Summaries
- Supplementary References

#### Supplementary Figures and Legends

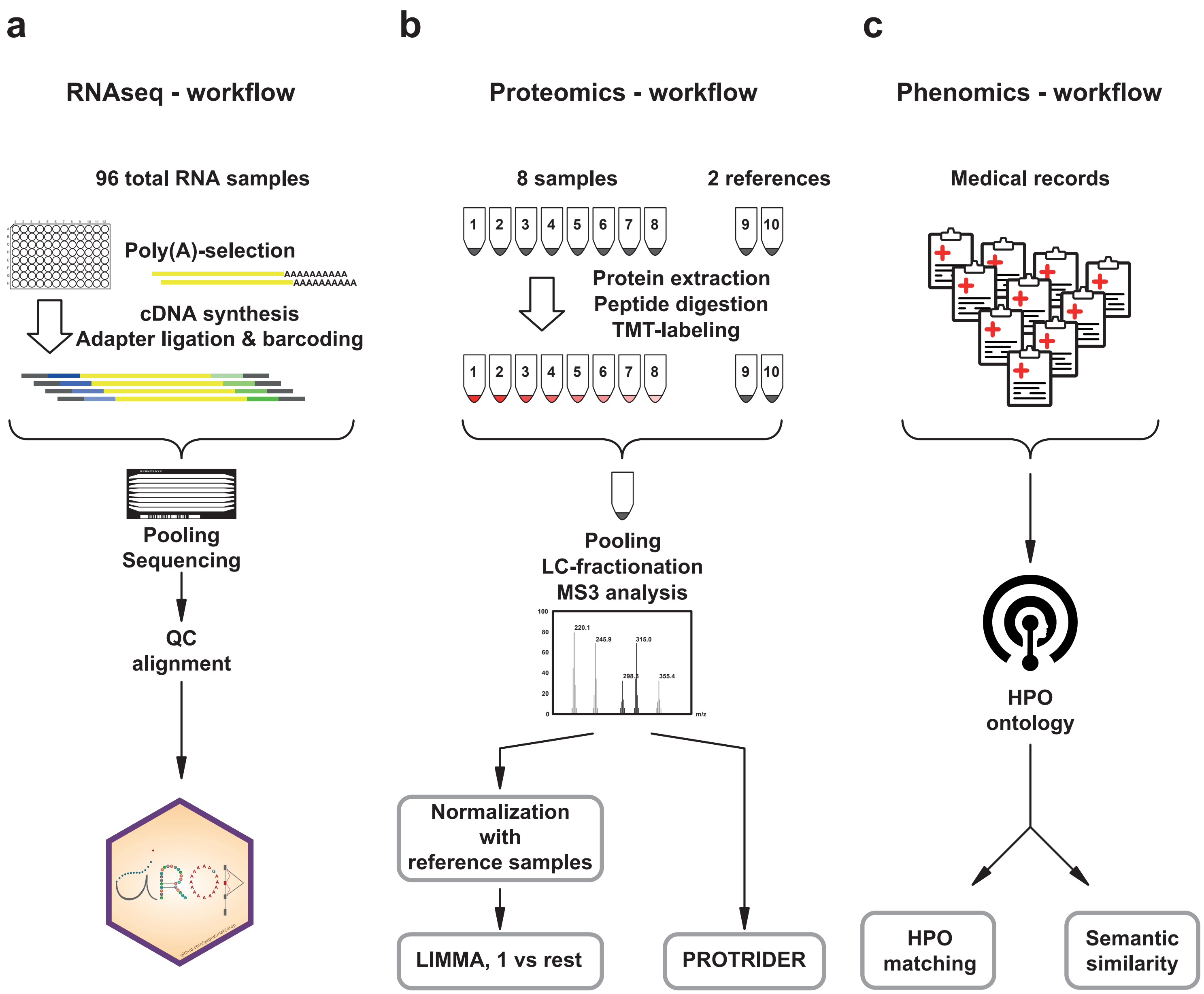
a
RNAseq - workflow
96 total RNA samples
Poly(A)-selection
AAAAAAAAAA
AAAAAAAAAA
cDNA synthesis
Adapter ligation & barcoding
Pooling
Sequencing
QC
alignment
b
Proteomics - workflow
8 samples
2 references
1 2 3 4 5 6 7 8
9 10
Protein extraction
Peptide digestion
TMT-labeling
1 2 3 4 5 6 7 8
9 10
Pooling
LC-fractionation
MS3 analysis
100
80
60
40
20
0
220.1
245.9
315.0
298.3
355.4
m/z
Normalization
with
reference samples
LIMMA, 1 vs rest
PROTRIDER
c
Phenomics - workflow
Medical records
HPO
ontology
HPO
matching
Semantic
similarity

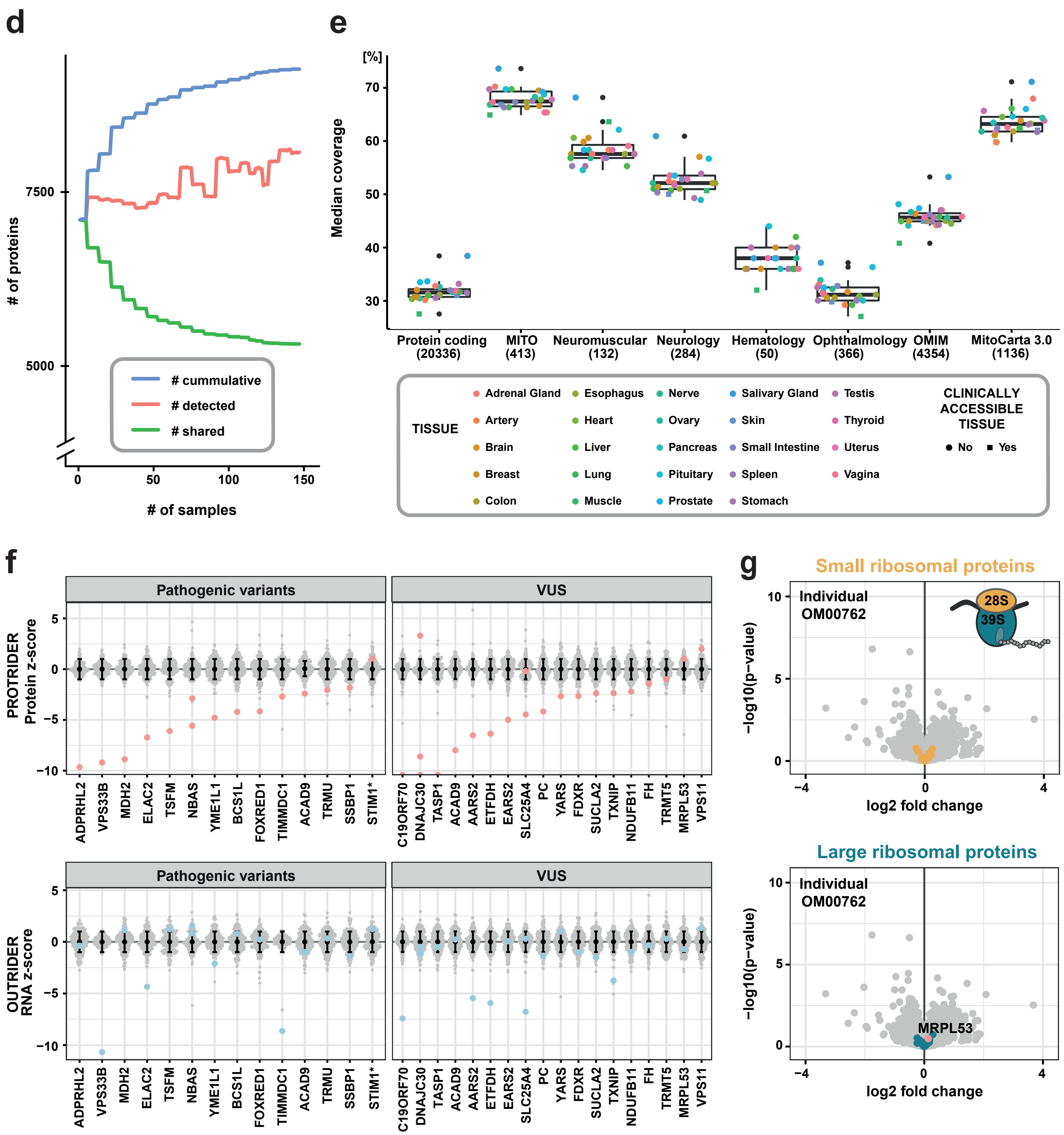
d
### of proteins
7500
5000
0
50
100
150
### of samples
### cummulative
### detected
### shared
e
[%]
Median coverage
70
60
50
40
30
Protein coding (20336)
MITO (413)
Neuromuscular (132)
Neurology (284)
Hematology (50)
Ophthalmology (366)
OMIM (4354)
MitoCarta 3.0 (1136)
TISSUE
Adrenal Gland
Artery
Brain
Breast
Colon
Esophagus
Heart
Liver
Lung
Muscle
Nerve
Ovary
Pancreas
Pituitary
Prostate
Salivary Gland
Skin
Small Intestine
Spleen
Stomach
Testis
Thyroid
Uterus
Vagina
CLINICALLY ACCESSIBLE TISSUE
No
Yes
f
Pathogenic variants
VUS
PROTRIDER Protein z-score
OUTRIDER RNA z-score
5
0
−5
−10
ADPRHL2
VPS33B
MDH2
ELAC2
TSFM
NBAS
YME1L1
BCS1L
FOXRED1
TIMMDC1
ACAD9
TRMU
SSBP1
STIM1*
C19ORF70
DNAJC30
TASP1
ACAD9
AARS2
ETFDH
EARS2
SLC25A4
PC
YARS
FDXR
SUCLA2
TXNIP
NDUFB11
FH
TRMT5
MRPL53
VPS11
g
Small ribosomal proteins
Individual OM00762
28S
39S
−log10(p−value)
10
5
0
−4
0
4
log2 fold change
Large ribosomal proteins
Individual OM00762
MRPL53

**Supplementary Fig. 1: Transcriptomic, proteomic, and phenomic workflows and the detection of proteins and outliers. a,** RNA-seq analysis of patient-derived fibroblasts was performed by strand-specific mRNA sequencing in batches of 96 samples. Discarding lowly expressed genes, approximately 12,000 transcripts were detectable (total 15,652, median 11,432). Data analysis was performed in the DROP pipeline (see **Methods**). **b,** Proteomic analysis of patient-derived fibroblasts was performed by tandem mass tag (TMT) labelled proteomics. Eight patient samples and two reference samples were processed in parallel. Peptides were tagged sample-wise prior to pooling, liquid chromatography (LC) fractionation, and analysis by mass spectrometry (MS3). The reference sample, in duplicate, was included for inter-batch normalization. Approximately 8,000 proteins were quantified per sample (total 9,264, median 7,720). Data analysis was performed by both limma and PROTRIDER (see **Methods**). **c,** Phenomic workflow from extraction of patient phenotype data from the medical records, reported as human phenotype ontology (HPO) terms, to calculation of the symmetric semantic similarity between the patient's phenotype and the disease-gene-associated phenotype reported in OMIM (https://omim.org) and determination of a phenotype match to the disease-gene-associated phenotype (see **Methods**). **d,** Number of detected proteins cumulatively (blue), per sample (red), and shared across all samples (green) in our dataset. **e,** Mean coverage of proteins encoded by all protein coding genes, a selection of disease genes for various disease groups, and all OMIM disease genes by tissue as per the publically available GTEx proteomic dataset (Jiang et al., 2020). The center line represents median, box limits correspond to the 25th and 75th percentiles, whiskers extend to the largest value no further than 1.5 * IQR from the box limits, values beyond the whiskers are outliers and plotted individually. **f,** Protein and RNA z-score distribution for disease-causing genes with known pathogenic variants (left panels) and prioritized variants of uncertain significance (VUS, right panels). The points appear in red/blue for cases known to harbour variants requiring validation and in green for novel cases diagnosed in our downstream systematic approach. Proteins and RNAs that were not detected are depicted on the y-axis at a z-score value of -10. **g,** Normal expression of MRPL53 and both the small and large mitoribosomal subunit in individual OM00762. Data are displayed as a gene-wise protein expression volcano plot of nominal ($-\log_{10}$) p-values against protein intensity $\log_2$ fold change.

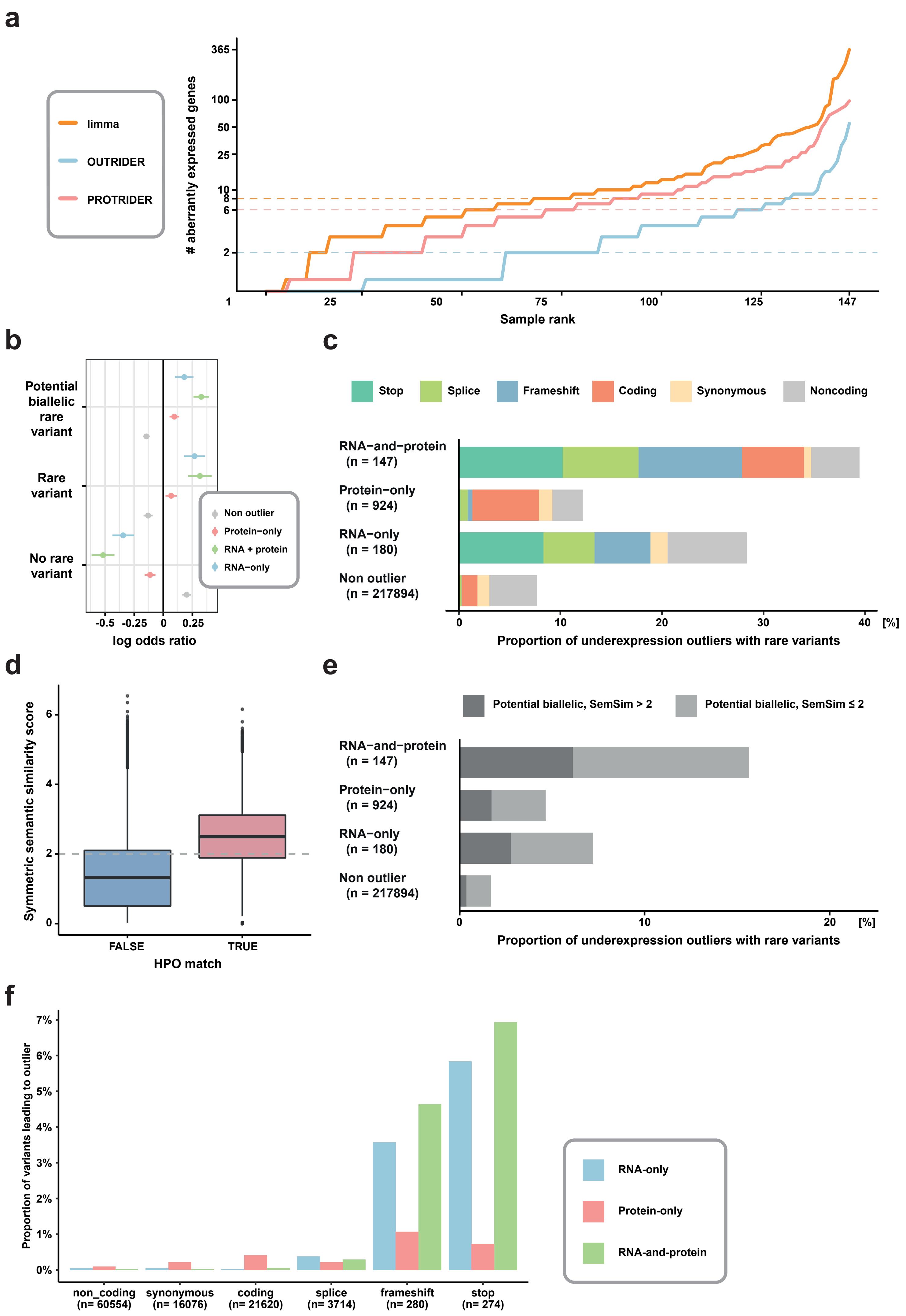
a
limma
OUTRIDER
PROTRIDER
### aberrantly expressed genes
365
100
50
25
10
8
6
2
1
25
50
75
100
125
147
Sample rank
b
Potential biallelic rare variant
Rare variant
No rare variant
Non outlier
Protein−only
RNA + protein
RNA−only
-0.5
-0.25
0
0.25
log odds ratio
c
Stop
Splice
Frameshift
Coding
Synonymous
Noncoding
RNA−and−protein (n = 147)
Protein−only (n = 924)
RNA−only (n = 180)
Non outlier (n = 217894)
0
10
20
30
40
[%]
Proportion of underexpression outliers with rare variants
d
Symmetric semantic similarity score
6
4
2
0
FALSE
TRUE
HPO match
e
Potential biallelic, SemSim > 2
Potential biallelic, SemSim ≤ 2
RNA−and−protein (n = 147)
Protein−only (n = 924)
RNA−only (n = 180)
Non outlier (n = 217894)
0
10
20
[%]
Proportion of underexpression outliers with rare variants
f
Proportion of variants leading to outlier
7%
6%
5%
4%
3%
2%
1%
0%
non_coding (n= 60554)
synonymous (n= 16076)
coding (n= 21620)
splice (n= 3714)
frameshift (n= 280)
stop (n= 274)
RNA-only
Protein-only
RNA-and-protein

**Supplementary Fig. 2: The association of RNA and protein outliers with genetic variants and phenotype. a,** Systematic calling of expression outliers over the entire dataset of 143 patients and two healthy controls identifying a median of two aberrantly expressed transcripts (OUTRIDER) and either six (PROTRIDER) or eight (limma) aberrantly expressed proteins per sample. **b,** Enrichment analysis for rare potentially biallelic, rare single, and frequent variants in all classes of outliers. Data are displayed as a log odds ratio with the 95% confidence interval. **c,** Proportion of underexpression outliers with rare variants detected by WES of three different outlier classes (RNA-only, protein-only, and RNA-and-protein) in comparison to non-outliers. 40% of RNA-and-protein outliers, 28% of RNA-only outliers, and 12% of protein-only outliers contain at least one rare variant in the corresponding gene, in comparison to 8% in non-outliers. **d,** Proportion of rare variants in a given category that led to an outlier across outlier classes. **e,** Semantic similarity calculated from patient HPO terms in patients with a phenotype match according to the method defined by Fresard et al., 2019. Semantic similarity threshold of ≥2 determined to be equivalent to a phenotype match. The center line represents median, box limits correspond to the 25th and 75th percentiles, whiskers extend to the largest value no further than 1.5 * IQR from the box limits, values beyond the whiskers are outliers and plotted individually. **f,** Proportion of underexpression outliers with rare potentially biallelic variants detected by WES of three different outlier classes (RNA-only, protein-only, and RNA-and-protein) in comparison to non-outliers stratified by semantic similarity score of ≥2 (phenotype match) and with a semantic similarity score of <2 or missing (no phenotype match).

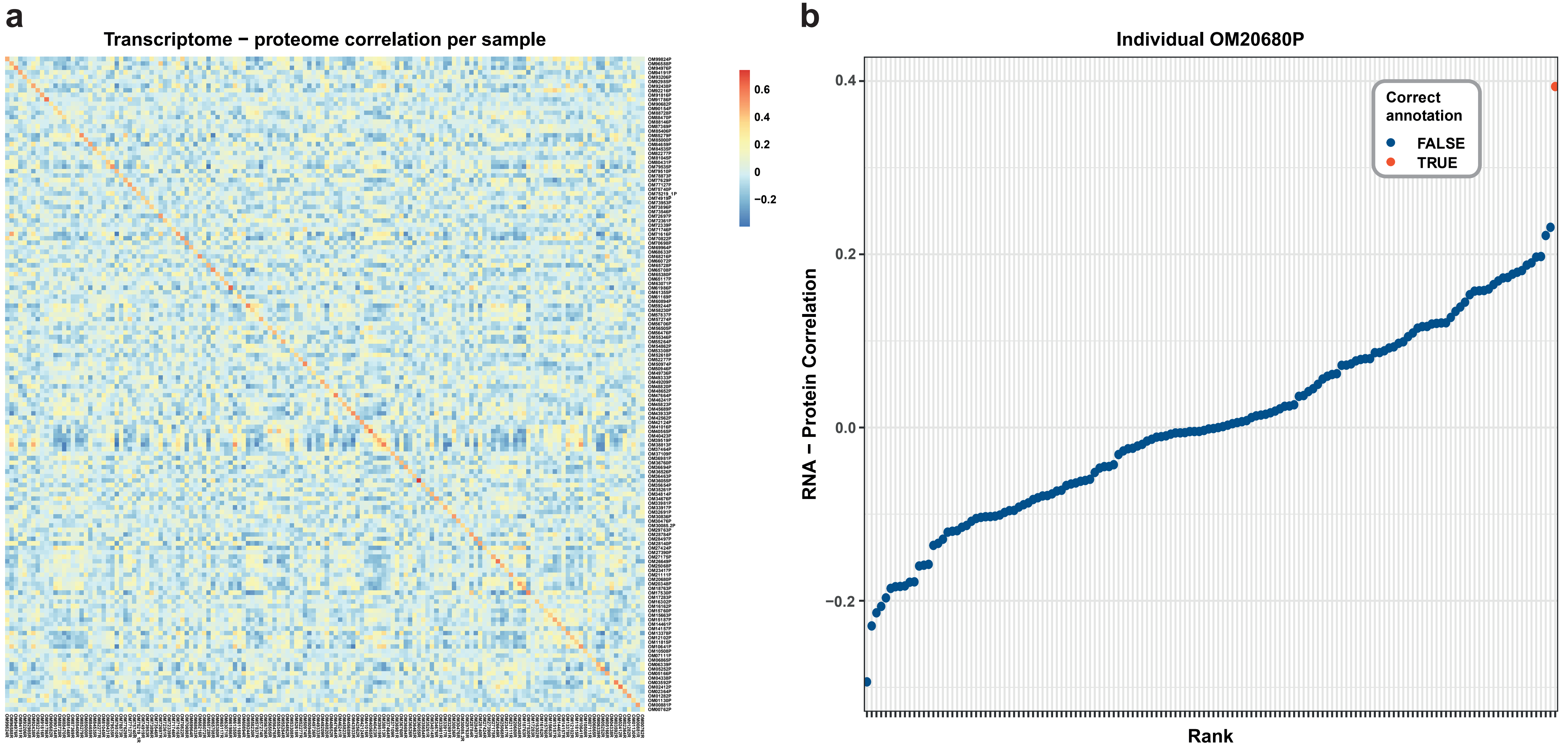
a
Transcriptome − proteome correlation per sample
0.6
0.4
0.2
0
−0.2
b
Individual OM20680P
RNA − Protein Correlation
0.4
0.2
0.0
−0.2
Rank
Correct annotation
FALSE
TRUE

**Supplementary Fig. 3: RNA-seq-proteomics matching. a,** Heatmap of sample-wise transcriptome-proteome Spearman rank correlation. Red indicates positive correlation, blue indicates negative correlation. **b,** Example of Spearman rank correlation for the proteome sample OM35261 to all transcriptomes, confirming the corresponding transcriptome according to the sample annotation (red) to demonstrate the highest correlation coefficient.

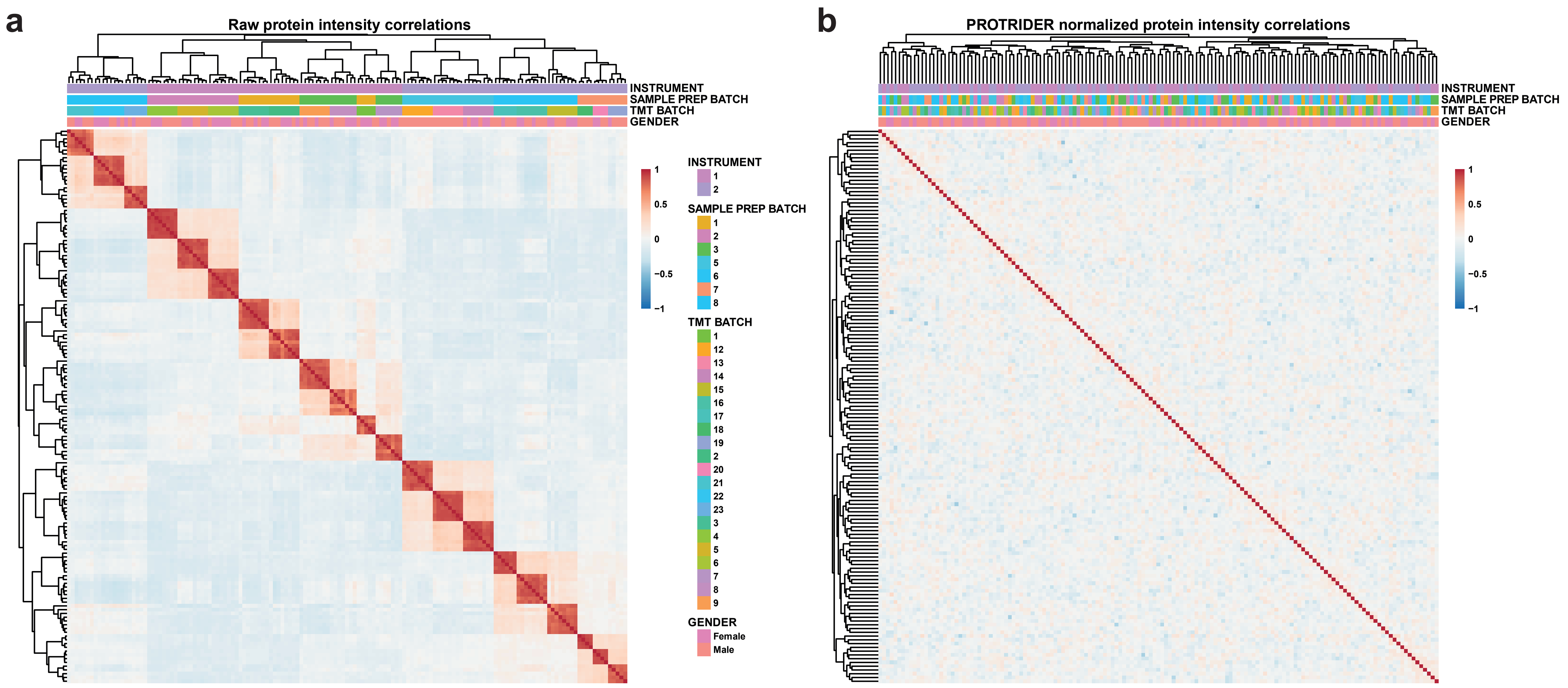
a
Raw protein intensity correlations
INSTRUMENT
SAMPLE PREP BATCH
TMT BATCH
GENDER
1
0.5
0
−0.5
−1
INSTRUMENT
1
2
SAMPLE PREP BATCH
1
2
3
5
6
7
8
TMT BATCH
1
12
13
14
15
16
17
18
19
2
20
21
22
23
3
4
5
6
7
8
9
GENDER
Female
Male
b
PROTRIDER normalized protein intensity correlations
INSTRUMENT
SAMPLE PREP BATCH
TMT BATCH
GENDER
1
0.5
0
−0.5
−1

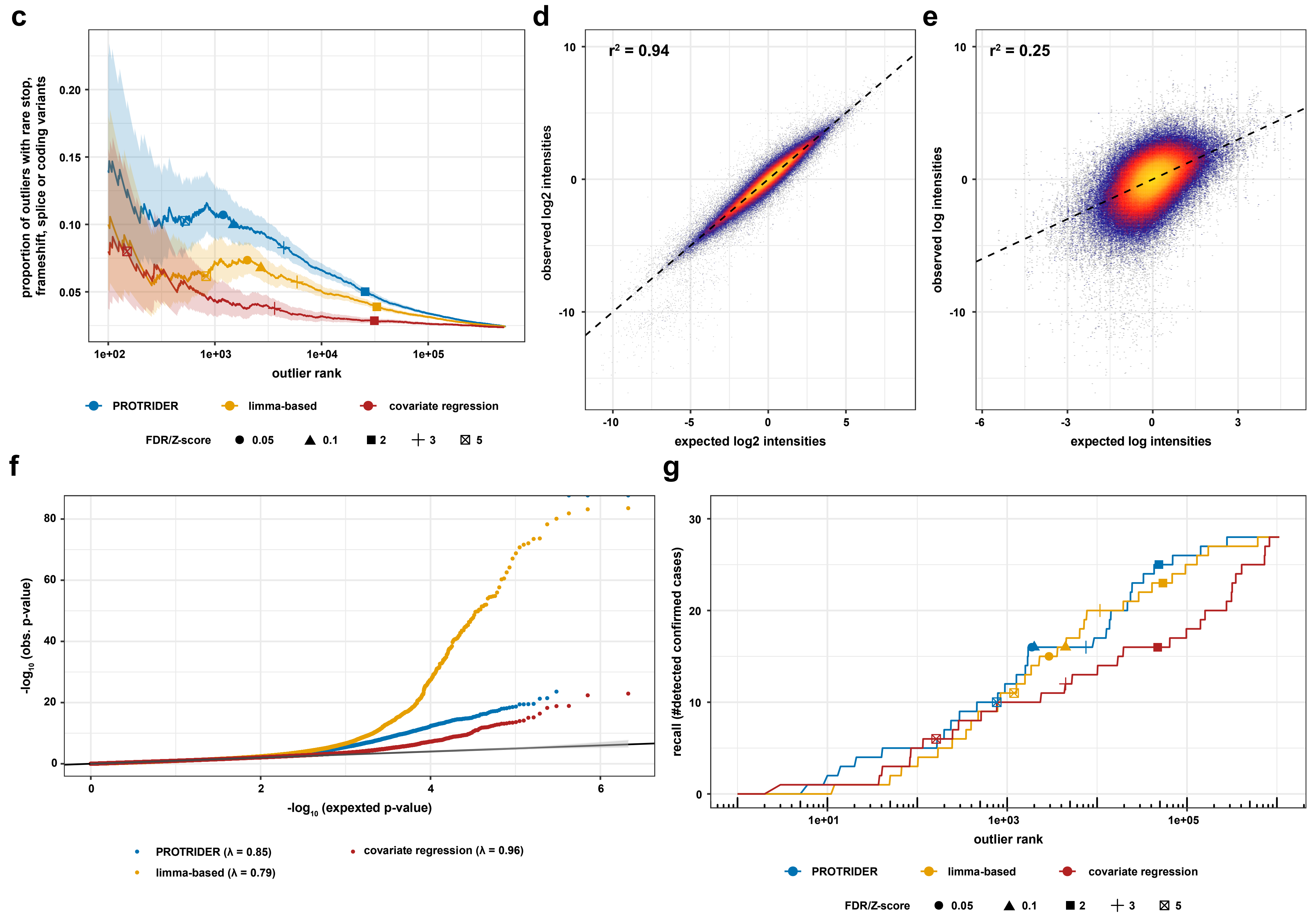
c
proportion of outliers with rare stop, frameshift, splice or coding variants
0.05
0.10
0.15
0.20
1e+02
1e+03
1e+04
1e+05
outlier rank
PROTRIDER
limma-based
covariate regression
FDR/Z-score
0.05
0.1
2
3
5
d
r² = 0.94
observed log2 intensities
expected log2 intensities
-10
-5
0
5
10
e
r² = 0.25
observed log intensities
expected log intensities
-6
-3
0
3
-10
0
10
f
-log10 (obs. p-value)
-log10 (expexted p-value)
0
20
40
60
80
0
2
4
6
PROTRIDER (λ = 0.85)
limma-based (λ = 0.79)
covariate regression (λ = 0.96)
g
recall (#detected confirmed cases)
0
10
20
30
1e+01
1e+03
1e+05
outlier rank
PROTRIDER
limma-based
covariate regression
FDR/Z-score
0.05
0.1
2
3
5

**Supplementary Fig. 4: Comparison of PROTRIDER with alternative approaches for the detection of aberrant protein expression.** Correlation matrix of samples (rows and columns) on **a,** raw log-transformed and protein-centered intensity values and **b,** PROTRIDER normalized log-transformed and protein-centered intensity values. Red indicates positive correlation, blue indicates negative correlation. Colored horizontal tracks display instrument, sample preparation batch, and TMT proteomic batch, and gender of the samples. Two different instruments have been used for the analysis, three batches were processed in parallel indicated by the sample preparation batch, and the samples have been analysed in 23 TMT batches. The dendrogram represents the sample-wise clustering. **c,** Proportion of outlier genes containing a rare (MAF < 0.01) stop, splice, frameshift or coding variant against the ranking of outliers for PROTRIDER (blue), the limma-based approach (yellow) and z-scores on the residuals of a regression of covariates (red). Shapes indicate FDR cutoffs of 0.05 and 0.1 as well as z-score cutoffs of 2, 3, and 5. **d-e,** Observed $log_2$ intensities centered per protein (y axis) are plotted against predicted $log_2$ insensities (x axis, also centered per protein) for **d,** PROTRIDER and **e,** the covariate regression approach. The color indicates the density of the data points with yellow corresponding to high density and blue to low density. $r^2$ values describing the proportion of variance explained are shown in the top left corner. **f,** Global quantile-quantile plot of observed negative log-transformed p-values (y-axis) against expected p-values (x-axis) for PROTRIDER (blue), the limma-based approach (yellow) and z-scores on the residuals of a regression of covariates (red). Under the null hypothesis, the data are expected to lie along the diagonal (95% confidence bands shown in gray). **g,** Recall (number of detected confirmed pathogenic outliers, n=28, y axis) is plotted against the ranking of nominal p-values of all protein-sample pairs (x axis) for PROTRIDER (blue), the limma-based approach (yellow) and z-scores on the residuals of a regression of covariates (red). Shapes indicate FDR cutoffs of 0.05 and 0.1 as well as z-score cutoffs of 2, 3 and 5.

#### Case Summaries

##### Validation cohort

##### Case 1, Sample ID: OM72697, Causal gene: *DNAJC30*

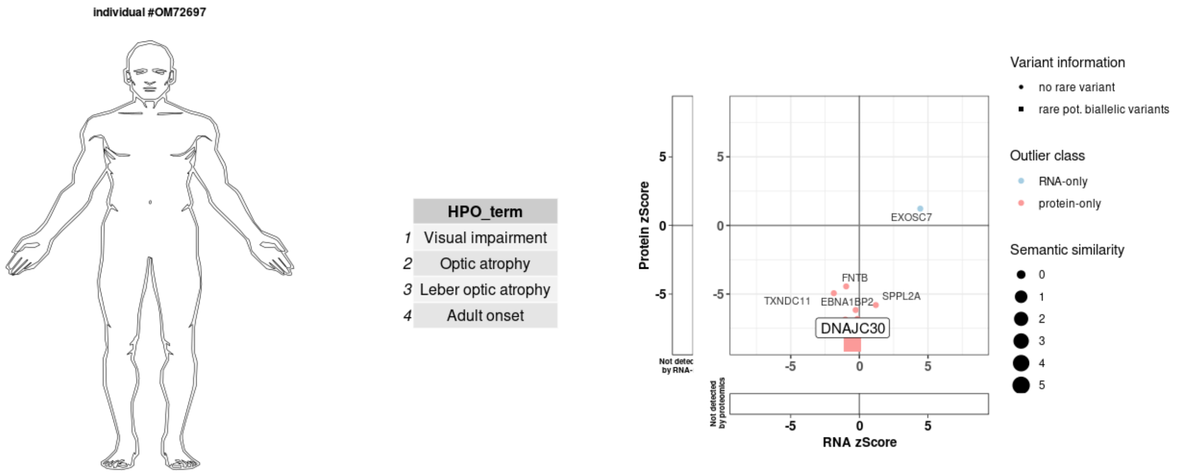


Adult male diagnosed with homozygous missense variant (c.[152A>G], p.[Tyr51Cys]) in *DNAJC30*. Phenotype of the patient matched the published phenotype of the causal gene (Semantic similarity score = 5.95). DNAJC30 was a significant protein underexpression outlier (FC: 0.01; p-value: 8.72*10-18; z-score: -8.59). The protein was detected in the TMT-batch but not in this sample, therefore the missing value was imputed as described in the methods.

**Case 2, Sample ID: OM43933, Causal gene: *DNAJC30***

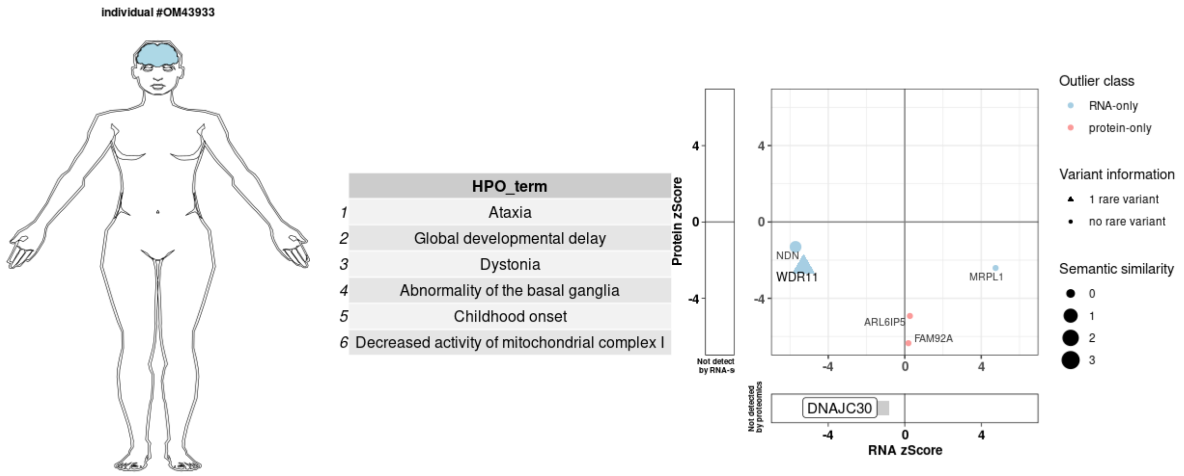


Infant female diagnosed with homozygous missense variant (c.[152A>G], p.[Tyr51Cys]) in *DNAJC30*. Phenotype of the patient matched the published phenotype of the causal gene (Semantic similarity score = 6.54). DNAJC30 was a significant protein underexpression outlier (FC: 0.15; p-value: 1.66*10-15; z-score: -7.96). The protein was detected in the replicate sample grown in galactose medium.

**Case 3, Sample ID: OM88470, Causal gene: *PC***

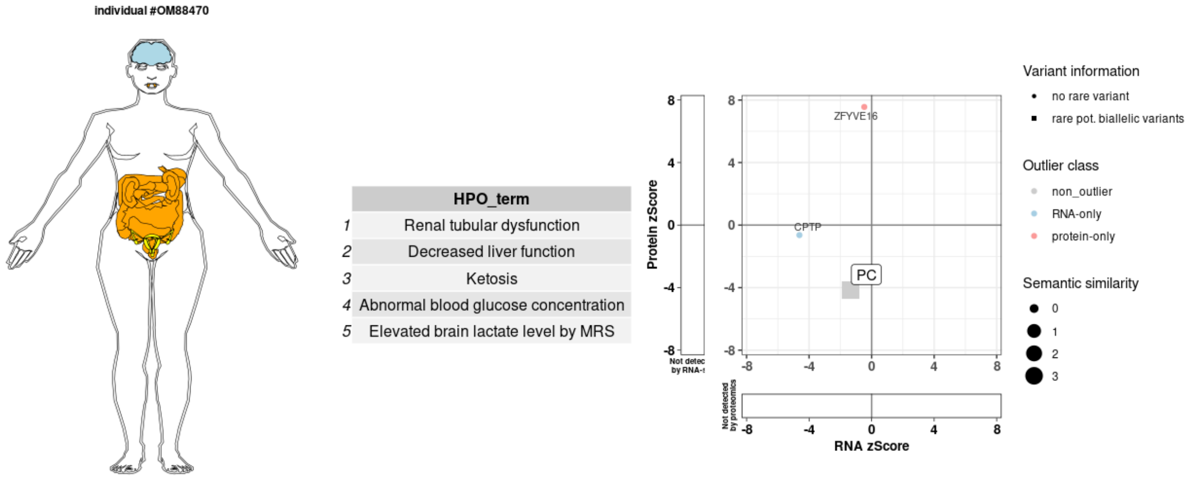


Paediatric female diagnosed with compound heterozygous missense variants (c.[1217G>A], p.[Arg406His]; c.[1891C>T], p.[Arg631Trp]) in *PC*. Phenotype of the patient matched the published phenotype of the causal gene (Semantic similarity score = 3.61). PC protein was underexpressed with nominal significance (FC: 0.52; p-value: 3.19*10-5; z-score: -4.16).

**Case 4, Sample ID: OM26649, Causal gene: *AARS2***

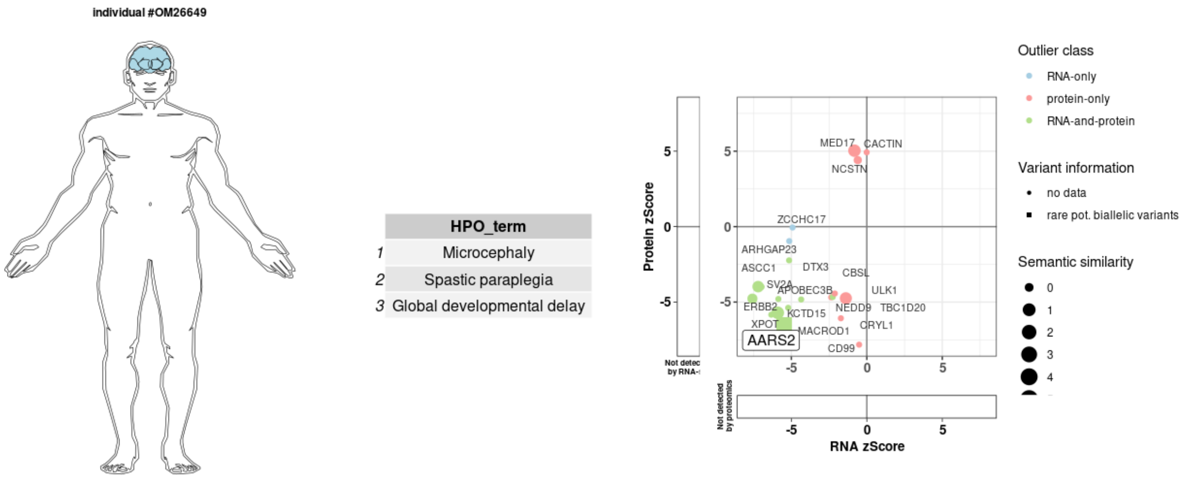


Paediatric male diagnosed with homozygous missense variant (c.[959C>T], p.[Ala320Val]) in *AARS2*. Phenotype of the patient matched the published phenotype of the causal gene (Semantic similarity score = 3.18). AARS2 was significantl RNA-and-protein underexpression outlier (RNA: FC: 0.54; p-value: 5.49*10-7; z-score: -5.46 / Protein: FC: 0.20; p-value: 7.91*10-11; z-score: -6.50).

**Case 5, Sample ID: OM70698, Causal gene: *TXNIP***

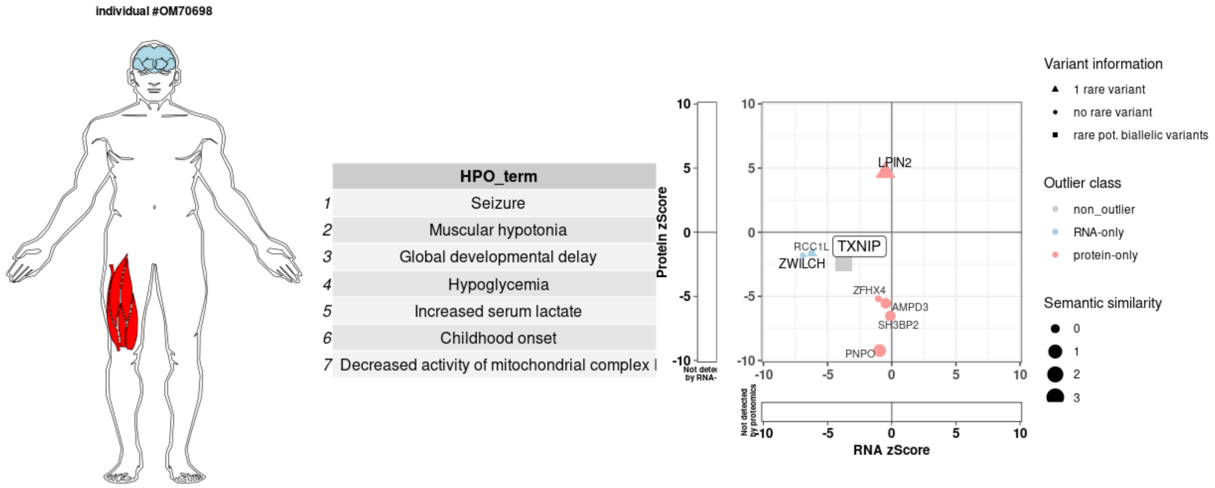


Paediatric male diagnosed with homozygous indel variant causing frameshift (c.[642_643insT], p.[Ile215Tyrfs*59]) in *TXNIP*. Phenotype of the patient matched the published phenotype of the causal gene (Semantic similarity score = 3.05). *TXNIP* was not published as a disease-associated gene when the case was initially investigated. Moreover, once reported, the phenotype similarity was questionable given that only one family has been published. TXNIP was underexpressed with nominal significance on both RNA and protein levels (RNA: FC: 0.18; p-value: 9.51*10-4; z-score: -3.75 / Protein: FC: 0.35; p-value: 1.87*10-2; z-score: -2.35).

**Case 6, Sample ID: OM50946, Causal gene: *NDUFB11***

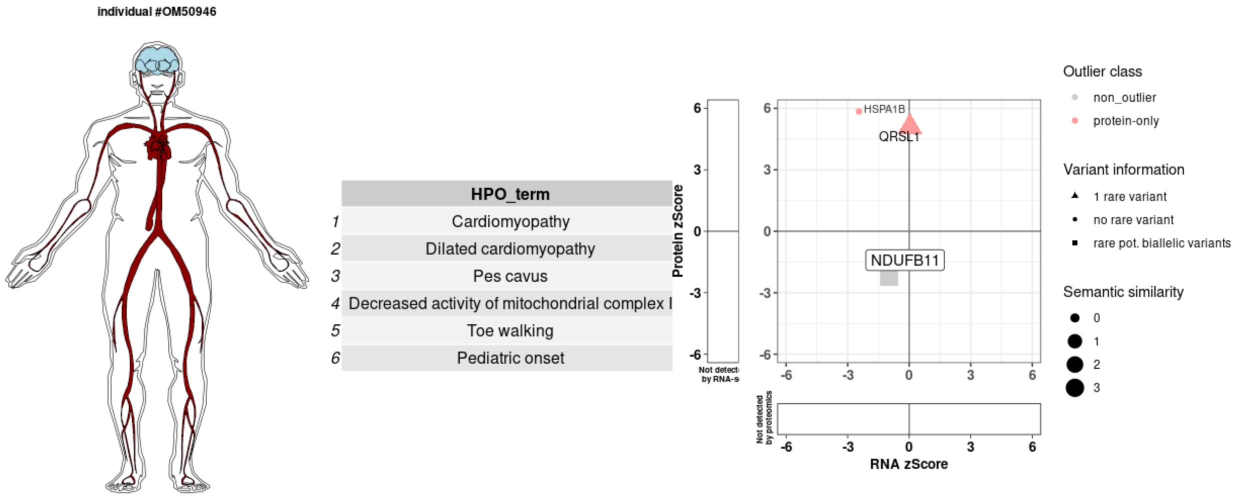


Paediatric male diagnosed with hemizygous missense variant (c.[379T>C], p.[Trp127Arg]) in *NDUFB11*. Phenotype of the patient matched the published phenotype of the causal gene (Semantic similarity score = 4.36). NDUFB11 protein was underexpressed with nominal significance (Protein: FC: 0.68; p-value: 2.78*10-2; z-score: -2.20). Furthermore a global reduction of respiratory chain complex I was observed.

#### Case 7, Sample ID: OM05252, Causal gene: *EARS2*

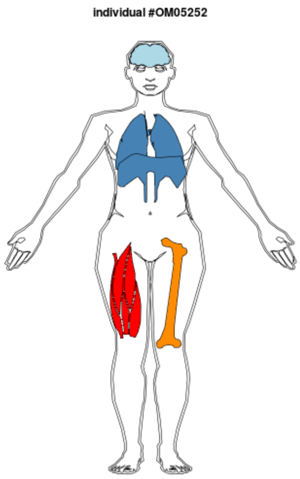


| | HPO_term |
|---|---|
| 1 | Microcephaly |
| 2 | Muscular hypotonia |
| 3 | Agenesis of corpus callosum |
| 4 | Encephalopathy |
| 5 | Abnormality of coagulation |
| 6 | Respiratory distress |
| 7 | Rhabdomyolysis |
| 8 | Elevated serum creatine kinase |
| 9 | Abnormal circulating alanine concentration |
| 10 | Elevated brain lactate level by MRS |

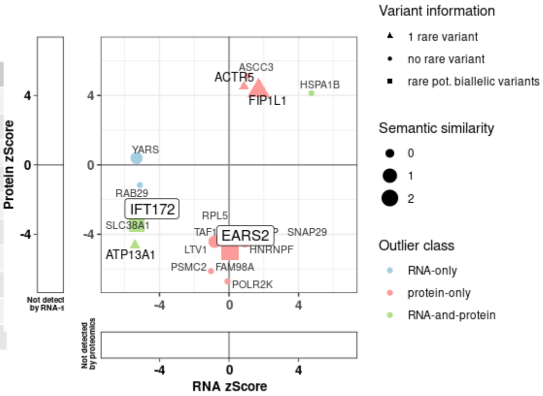


Paediatric female diagnosed with homozygous missense variant (c.[911C>A], p.[Pro304His]) in *EARS2*. Phenotype of the patient matched the published phenotype of the causal gene (Semantic similarity score = 2.96). EARS2 was a significant protein underexpression outlier (FC: 0.47; p-value: 6.57*10-7; z-score: -4.97).

**Case 8, Sample ID: OM77629, Causal gene: *ETFDH***

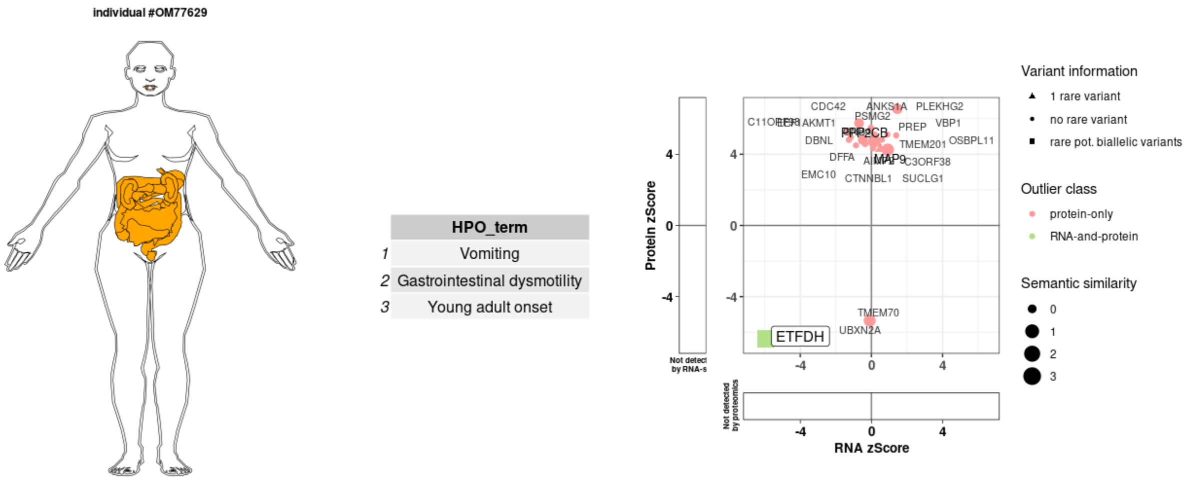


Adult female diagnosed with heterozygous indel variant (c.[685G>T; 687_688delAA; 691T>C];[=] p.[Ala229Ser; Thr230Ilefs*2; Phe231Val];[=]) in *ETFDH*. Phenotype of the patient matched the published phenotype of the causal gene (Semantic similarity score = 3.53). ETFDH was significant RNA-and-protein underexpression outlier (RNA: FC: 0.60; p-value: 2.71*10-8; z-score: -5.93 / Protein: FC: 0.48; p-value: 2.22*10-10; z-score: -6.34.

#### Case 9, Sample ID: OM23417, Causal gene: *FDXR*

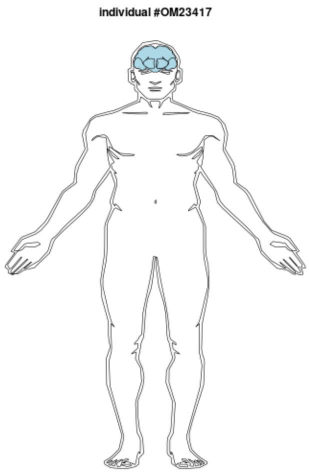


| | HPO_term |
|---|---|
| 1 | Sensorineural hearing impairment |
| 2 | Visual impairment |
| 3 | Seizure |
| 4 | Abnormality of the cerebral white matter |
| 5 | Neonatal onset |
| 6 | Neurodevelopmental abnormality |

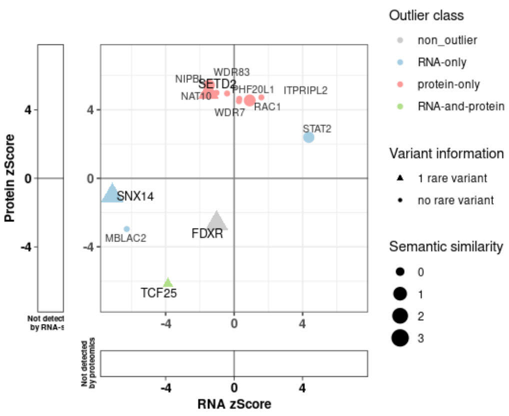


Neonatal male diagnosed with compound heterozygous missense variants (c.[683G>T], p.[Arg228Leu]; c.[1058G>A], p.[Cys353Tyr]) in *FDXR*. Phenotype of the patient matched the published phenotype of the causal gene (Semantic similarity score = 3.53). FDXR protein was underexpressed with nominal significance (FC: 0.63; p-value: 8.31*10-3; z-score: -2.64).

**Case 10, Sample ID: OM61355, Causal gene: *YARS***

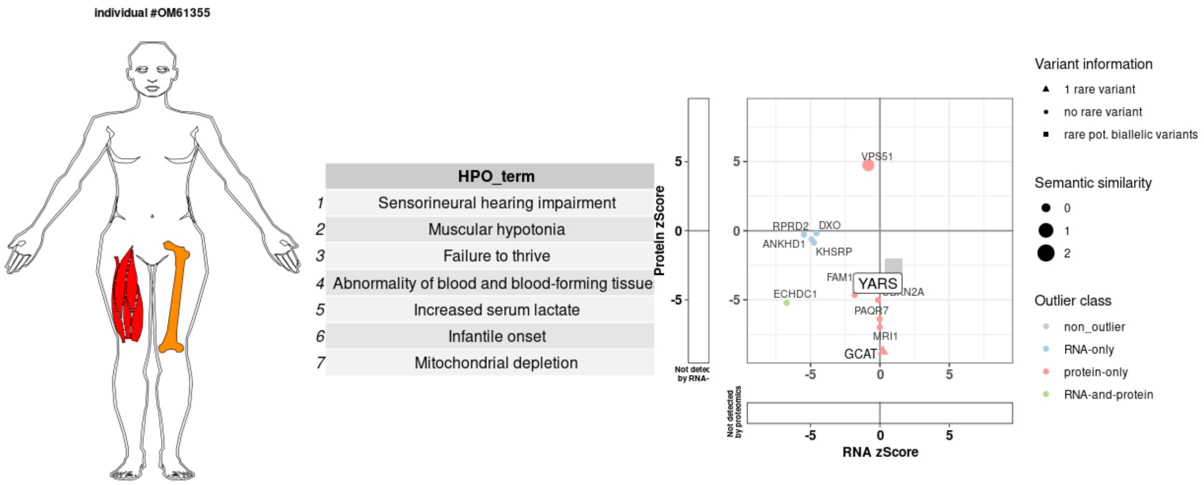


Infant female diagnosed with homozygous missense variant (c.[176T>C], p.[Ile59Thr]) in *YARS*. Phenotype of the patient matched the published phenotype of the causal gene (Semantic similarity score = 2.78). YARS protein was underexpressed with nominal significance (FC: 0.81; p-value: 7.87*10-3; z-score: -2.66).

**Case 11, Sample ID: OM65708, Causal gene: *ACAD9***

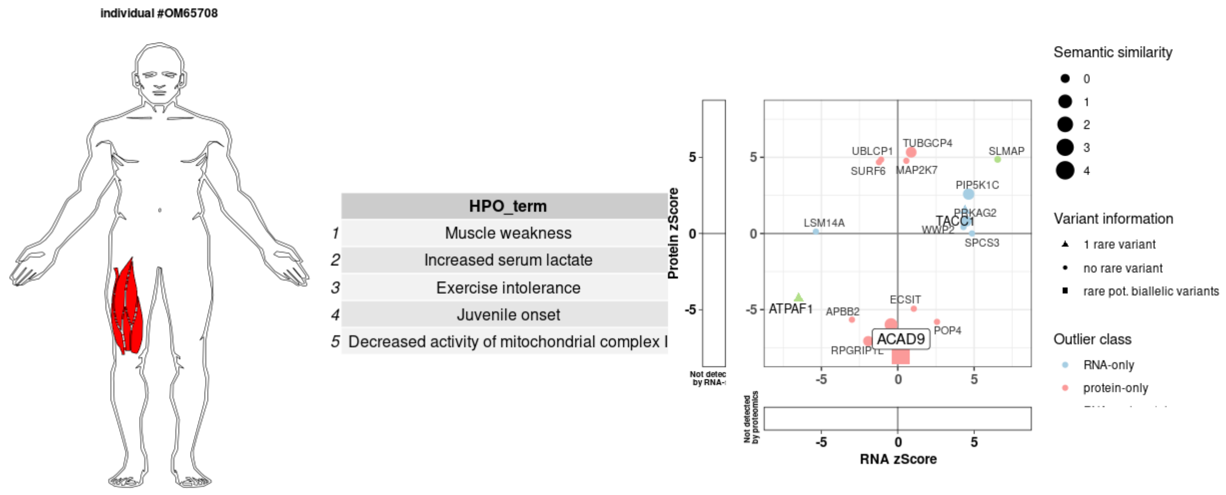


Paediatric male diagnosed compound heterozygous start loss and missense variants (c.[1A>G], p.[?]; c.[261T>G], p.[Ile87Met]) in *ACAD9*. Phenotype of the patient matched the published phenotype of the causal gene (Semantic similarity score = 4.37). ACAD9 was a significant protein underexpression outlier (FC: 0.29; p-value: 1.51*10-15; z-score: -7.98). ECSIT protein, direct interaction partner of ACAD9, also found significantly reduced (FC: 0.37; p-value: 7.89*10-7; z-score: -4.94).

**Case 12, Sample ID: OM27175, Causal gene: *FH***

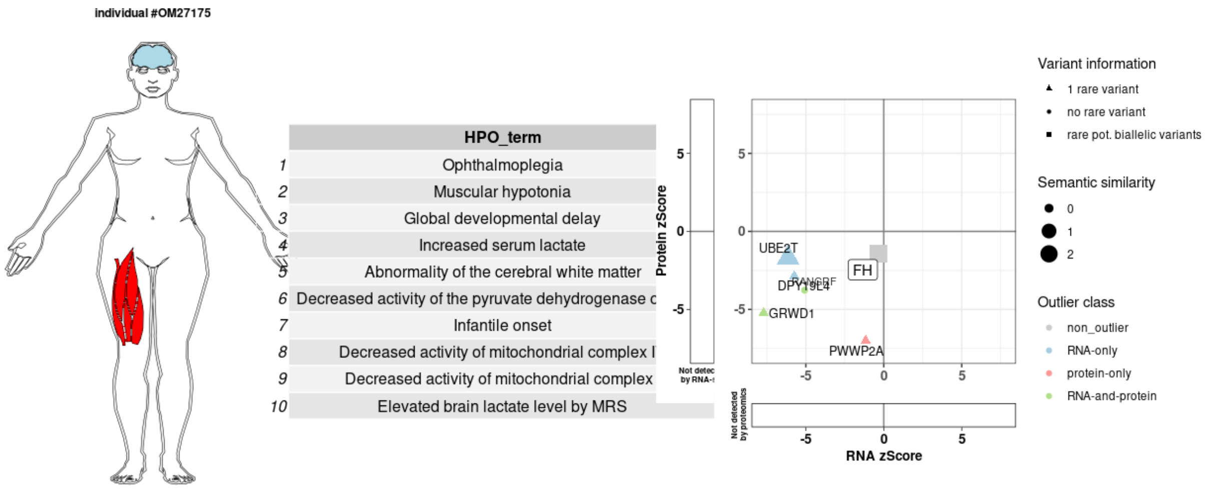


Infant female diagnosed with homozygous missense variant (c.[1048C>T], p.[Arg350Trp]) in *FH*. Phenotype of the patient matched the published phenotype of the causal gene (Semantic similarity score = 2.76). FH protein was underexpressed with nominal significance (FC: 0.88; p-value: 0.15; z-score: -1.44).

**Case 13, Sample ID: OM20348, Causal gene: *SUCLA2***

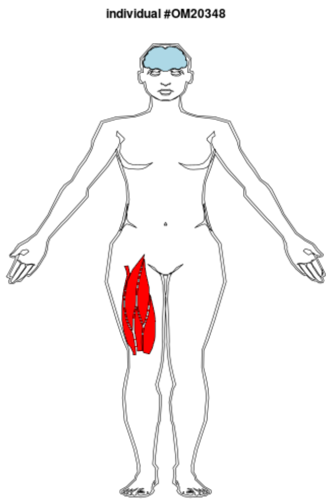


| | HPO_term |
|---|---|
| 1 | Microcephaly |
| 2 | Sensorineural hearing impairment |
| 3 | Delayed speech and language development |
| 4 | Intellectual disability |
| 5 | Seizure |
| 6 | Muscular hypotonia |
| 7 | Global developmental delay |
| 8 | Motor delay |
| 9 | Muscle weakness |
| 10 | Abnormality of the basal ganglia |
| 11 | Increased serum lactate |
| 12 | Developmental regression |
| 13 | Increased CSF lactate |
| 14 | Hyperalaninemia |
| 15 | 3-Methylglutaconic aciduria |
| 16 | Muscular dystrophy |
| 17 | Neonatal onset |
| 18 | Methylmalonic aciduria |

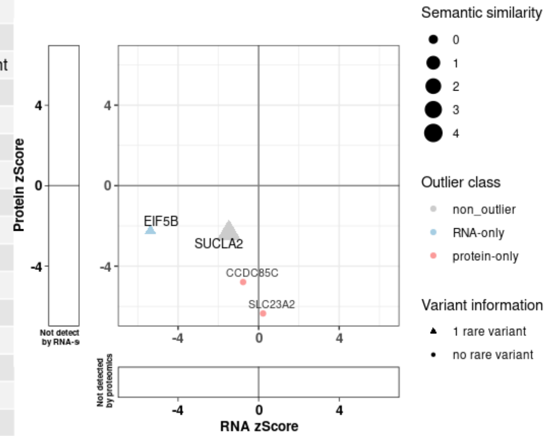


Infant female diagnosed with compound heterozygous missense variants (c.[670C>T], p.[Arg224Cys]; c.[805A>G], p.[Met269Val]) in *SUCLA2*. Phenotype of the patient matched the published phenotype of the causal gene (Semantic similarity score = 4.7). SUCLA2 protein was underexpressed with nominal significance (FC: 0.81; p-value: 1.77*10-2; z-score: -2.37). Furthermore a global reduction of Succinyl-CoA synthetase protein complex was observed.

**Case 14, Sample ID: OM02364, Causal gene: SLC25A4**

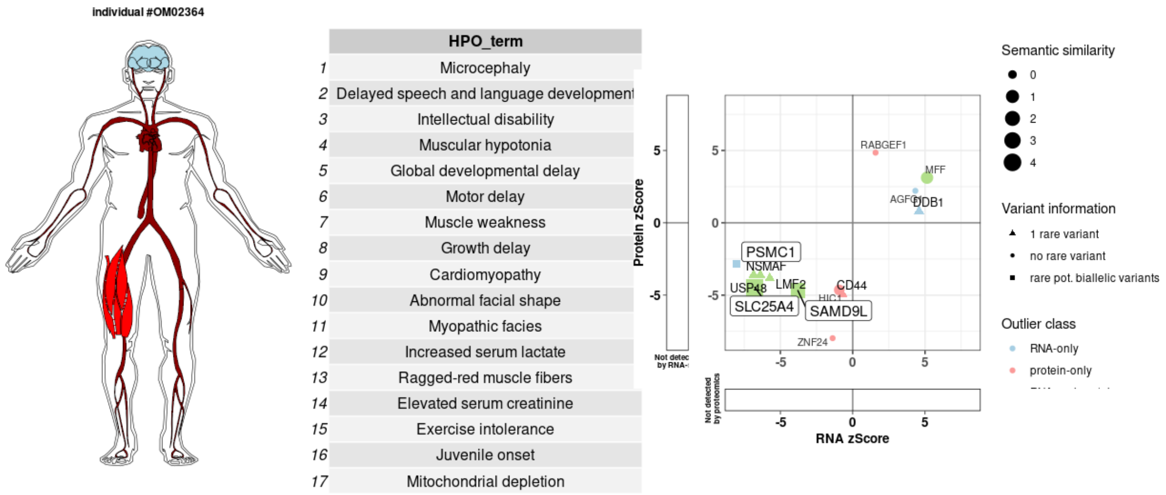


Paediatric male diagnosed with homozygous missense variant (c.[598G>A], p.[Gly200Arg]) in *SLC25A4*. Phenotype of the patient matched the published phenotype of the causal gene (Semantic similarity score = 4.29). SLC25A4 was a significant RNA-and-protein underexpression outlier (RNA: FC: 0.24; p-value: 6.39*10-9; z-score: -6.77 / Protein: FC: 0.35; p-value: 8.28*10-6; z-score: -4.46).

#### Case 21 - validated with RNA-seq, Sample ID: OM66072, Causal gene: *C19ORF70*

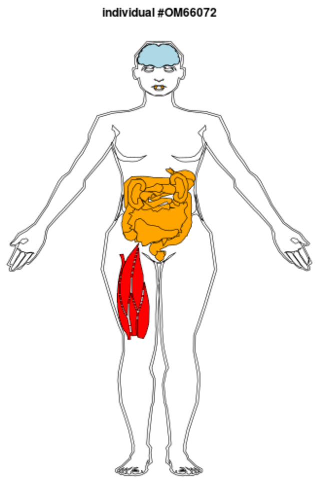


| | HPO_term |
|---|---|
| 1 | Muscular hypotonia |
| 2 | Global developmental delay |
| 3 | Abnormality of the liver |
| 4 | Decreased liver function |
| 5 | Intrauterine growth retardation |
| 6 | 3-Methylglutaconic aciduria |
| 7 | Antenatal onset |

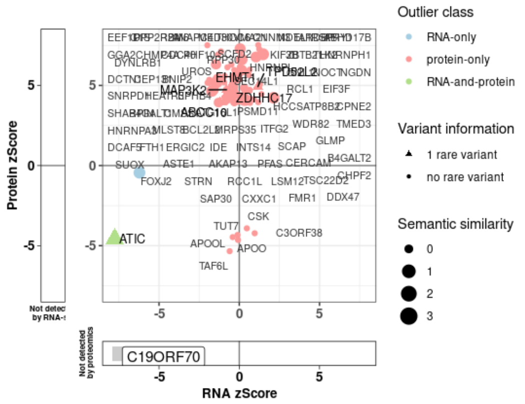


Paediatric female diagnosed with compound heterozygous frameshift and intronic variants (c.[143delT], p.[Val48Alafs*42]; c.[29+272G>C], p.[Phe11Trpfs*5]) in *c19orf70*. Phenotype of the patient matched the published phenotype of the causal gene (Semantic similarity score = 4.49). c19orf70 transcript was underexpressed with nominal significance and aberrant splicing was detected as well (RNA: FC: 0.59, p-value: 7.41*10-13; z-score: -7.43). Protein not detected in corresponding TMT-batch. Furthermore a global reduction of the MICOS complex was observed.

#### Discovery cohort

##### Case 1, Sample ID: OM91786, Causal gene: *LIG3*

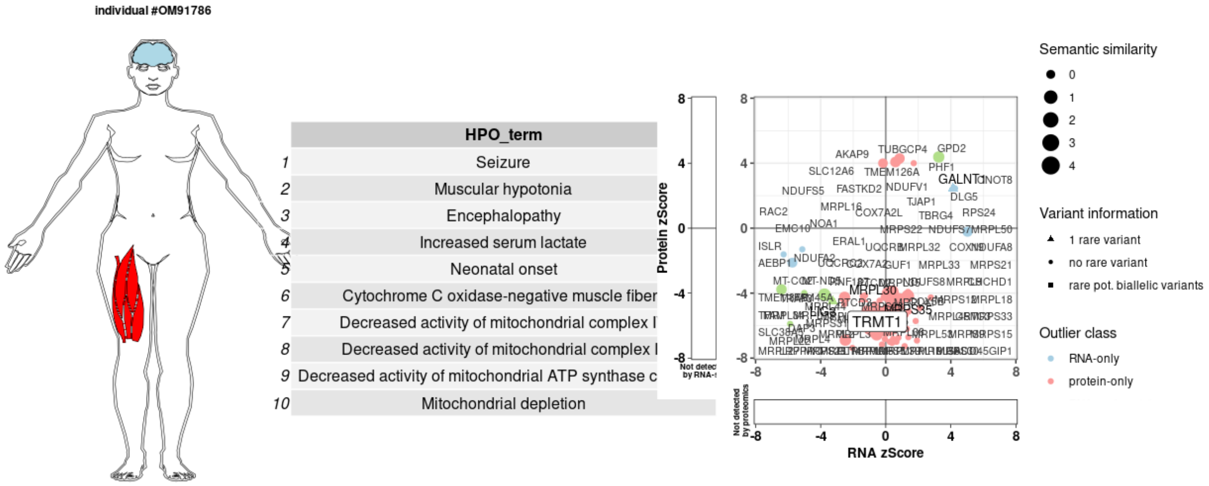


Infant female diagnosed with compound heterozygous nonsense and intronic variants (c.[86G>A], p.[Trp29*]; c.[1611+208G>A], p.[?]) in *LIG3*. LIG3 was a significant RNA-and-protein underexpression outlier (RNA: FC: 0.78; p-value: 0.0018; z-score: -3.26 / Protein: FC: 0.73; p-value: 6.94*10-6; z-score: -4.50). Intronic variant resulted in splice defect and NMD. Nonsense variant present in 70% of RNA reads, affecting only mitochondrial isoform. Defect in LIG3 caused significantly reduced expression of mtDNA encoded genes, explaining further 63 outliers. Furthermore a global reduction of multiple mitochondrial complexes including OXPHOS and mitochondrial ribosome was observed. While this manuscript was under review, *LIG3* was published as a novel mitochondrial disease-associated gene (Bonora et al., 2021).

**Case 2, Sample ID: OM57837, Causal gene: *MRPL38***

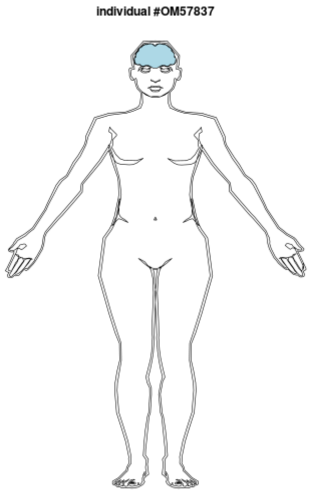


| | HPO_term |
|---|---|
| 1 | Intellectual disability |
| 2 | Seizure |
| 3 | Global developmental delay |
| 4 | Encephalopathy |
| 5 | Abnormality of the basal ganglia |
| 6 | Abnormality of brainstem morphology |
| 7 | Infantile onset |
| 8 | Decreased activity of mitochondrial complex I\ |
| 9 | Decreased activity of mitochondrial complex I |

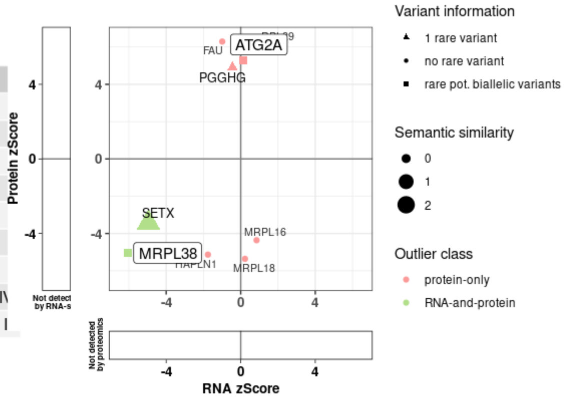


Infant female diagnosed with compound heterozygous 5'UTR-deletion and missense variants (c.[1-174del127], p.[?]; c.[770C>G], p.[Pro257Arg]) in *MRPL38*. MRPL38 was a significant RNA-and-protein underexpression outlier (RNA: FC: 0.66; p-value: 8.47*10-10; z-score: -6.05 // protein: FC: 0.66; p-value: 8.47*10-10; z-score: -6.05). Missense variant was present in 78% or RNA reads. Furthermore reduction of large mitochondrial ribosomal subunit was observed.

**Case 3, Sample ID: OM34814, Causal gene: *RDH11***

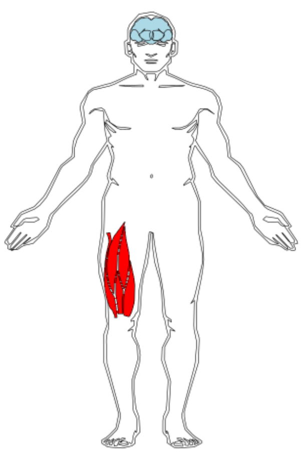


| | HPO_term |
|---|---|
| 1 | Microcephaly |
| 2 | Cataract |
| 3 | Seizure |
| 4 | Muscular hypotonia |
| 5 | Global developmental delay |
| 6 | Failure to thrive |
| 7 | Neurological speech impairment |
| 8 | Impairment of galactose metabolism |
| 9 | Hypergalactosemia |
| 10 | Pediatric onset |

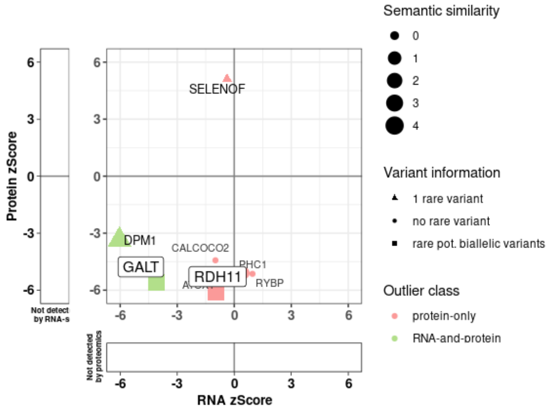


Paediatric male diagnosed with homozygous missense variant (c.[637A>C], p.[Thr213Pro]) in *RDH11*. Phenotype of the patient matched the published phenotype of the causal gene (Semantic similarity score = 2.96). RDH11 was a significant protein-only underexpression outlier (FC: 0.35; p-value: 9.89*10-10; z-score: -6.11). Furthermore reduction of RDH11-SELENOF protein complex was observed.

**Case 4, Sample ID: OM21111, Causal gene: *NFU1***

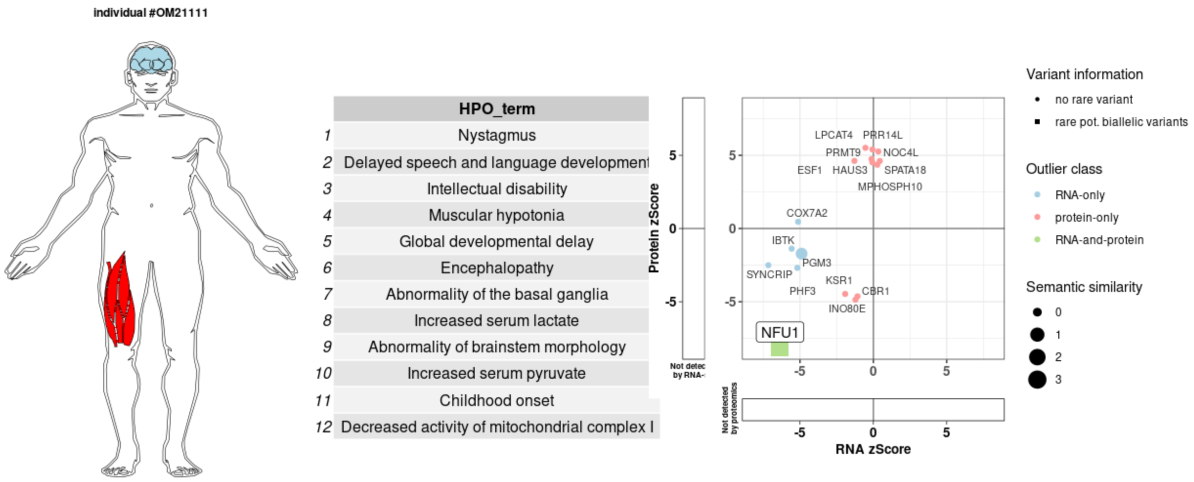


Infant male diagnosed with compound heterozygous missense and CNV variants (c.[362T>C], p.[Val121Ala]; c.[del exon5], p.[?]) in *NFU1*. Phenotype of the patient matched the published phenotype of the causal gene (Semantic similarity score = 3.26). NFU1 was a significant RNA-and-protein underexpression outlier. Whole exon deletion (exon 5 plus spanning region) resulted in significant RNA underexpression (FC: 0.63 p-value: 1.18*10-9; z-score: -6.36) plus monoallelic expressed missense variant resulted in protein underexpression (FC: 0.18; p-value: 4.00*10-16; z-score: -8.14).

#### Case 5, Sample ID: OM38813, Causal gene: *TIMMDC1*

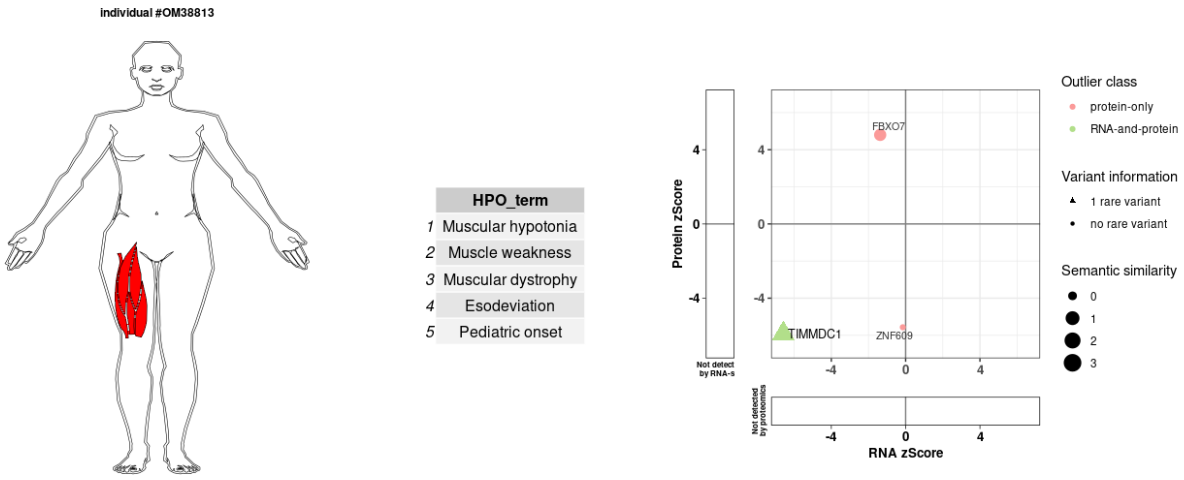


Paediatric male diagnosed with homozygous deep intronic variant (c.[597-1340A>G], p.[Gly199ins5*]) in *TIMMDC1*. Phenotype of the patient matched the published phenotype of the causal gene (Semantic similarity score = 3.53). Homozygous deep intronic variant caused splice defect (cryptic exon), resulting in frameshift and RNA-and-protein underexpression outlier (RNA: FC: 0.24; p-value: 3.66*10-11; z-score: -6.58 / Protein: FC: 0.27; p-value: 3.07*10-9; z-score: -5.93).

**Case 6, Sample ID: OM75740, Causal gene: *DARS2***

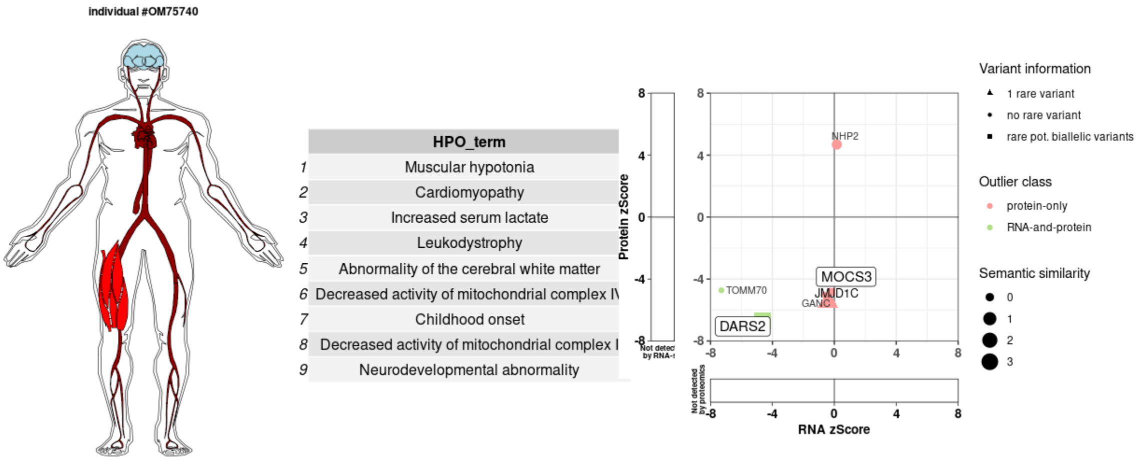


Infant male diagnosed with compound heterozygous nearsplice (1 rare + 1 frequent) and splice (rare) variants (c.[228-20T>C; 228-12C>G], p.[?]; c.[228-20T>C; 492+2T>C], p.[?]) in *DARS2*. Phenotype of the patient matched the published phenotype of the causal gene (Semantic similarity score = 4.03). DARS2 was a significant RNA-and-protein underexpression outlier (RNA: FC: 0.71; p-value: 5.95*10-5; z-score: -4.66 / Protein: FC: 0.37; p-value: 1.85*10-11; z-score: -6.72) due to partial skipping of exons 3 and 5.

**Case 7, Sample ID: OM06865, Causal gene: *EPG5***

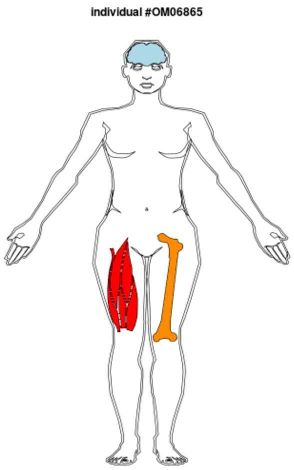


| | HPO_term |
|---|---|
| 1 | Ophthalmoplegia |
| 2 | Amblyopia |
| 3 | Optic atrophy |
| 4 | Seizure |
| 5 | Muscular hypotonia |
| 6 | Global developmental delay |
| 7 | Agenesis of corpus callosum |
| 8 | Encephalopathy |
| 9 | Muscle weakness |
| 10 | Hip dysplasia |
| 11 | Neutropenia |
| 12 | Hypoplasia of the corpus callosum |
| 13 | Scoliosis |
| 14 | Elevated hepatic transaminase |
| 15 | Ragged-red muscle fibers |
| 16 | Elevated serum creatine kinase |
| 17 | CNS demyelination |
| 18 | Decreased activity of mitochondrial complex I |
| 19 | Brain atrophy |
| 20 | Pediatric onset |

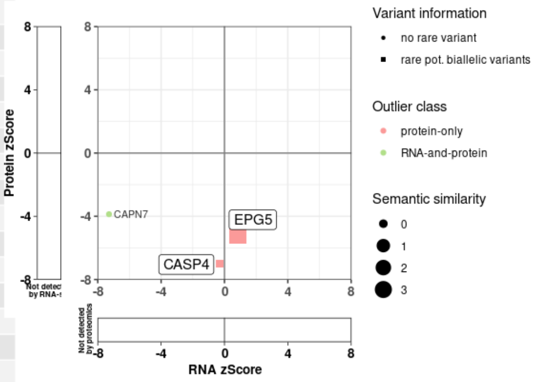


Paediatric female diagnosed with homozygous missense variant (c.[5548T>C], p.[Cys1850Arg]) in *EPG5*. Phenotype of the patient matched the published phenotype of the causal gene (Semantic similarity score = 3.73). EPG5 was a significant protein-only underexpression outlier (FC: 0.42; p-value: 2.13*10-7; z-score: -5.19).

#### Case 8, Sample ID: OM56706, Causal gene: *GMPPB*

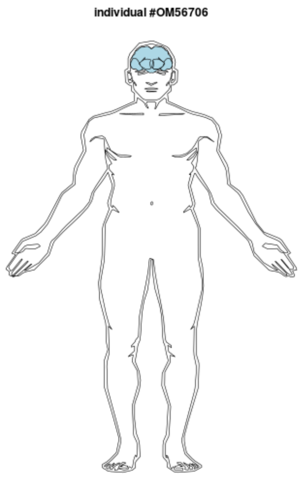


| | HPO_term |
|---|---|
| 1 | Optic atrophy |
| 2 | Seizure |
| 3 | Cerebellar atrophy |
| 4 | Hypoplasia of the corpus callosum |
| 5 | Cerebral cortical atrophy |
| 6 | Intellectual disability, profound |
| 7 | Cerebellar vermis atrophy |
| 8 | Intellectual disability, severe |

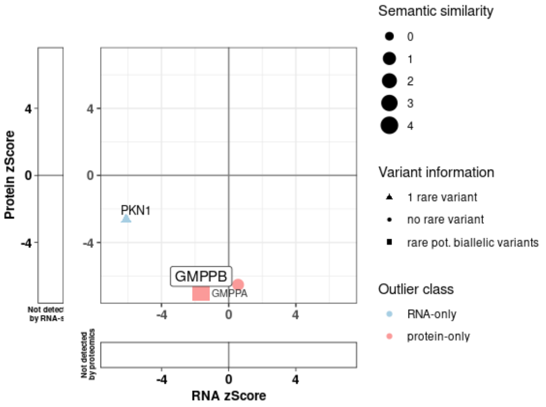


Paediatric male diagnosed with homozygous 5 bp duplication (c.[1016insAAGGC], p.[Tyr340Argfs*13]) in *GMPPB*. Phenotype of the patient matched the published phenotype of the causal gene (Semantic similarity score = 4.7). GMPPB was a significant protein-only underexpression outlier (Protein: FC: 0.40; p-value: 3.74*10-12; z-score: -6.95). Indel is intronic in the muscle-specific transcript isoform, resulting in a mild splice defect and exonic in brain-specific isoform leading to a frameshift in keeping with the patient presenting with neurological symptoms. Furthermore GMPPA protein, direct interaction partner of GMPPB, was also found as a significant protein-only underexpression outlier (Protein: FC: 0.48; p-value: 6.97*10-11; z-score: -6.52).

**Case 9, Sample ID: OM27390, Causal gene: *MORC2***

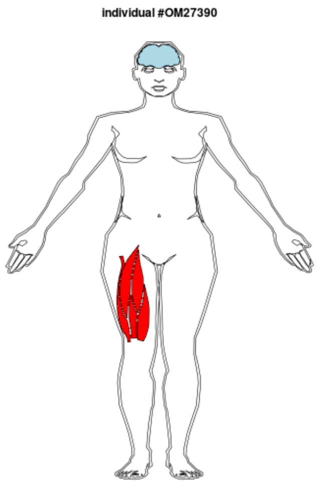


| | HPO_term |
|---|---|
| 1 | Nystagmus |
| 2 | Seizure |
| 3 | Muscular hypotonia |
| 4 | Global developmental delay |
| 5 | Encephalopathy |
| 6 | Failure to thrive |
| 7 | Abnormality of the basal ganglia |
| 8 | Increased serum lactate |
| 9 | Abnormality of brainstem morphology |
| 10 | Increased CSF lactate |
| 11 | Increased serum pyruvate |
| 12 | Infantile onset |
| 13 | Decreased activity of mitochondrial complex I |

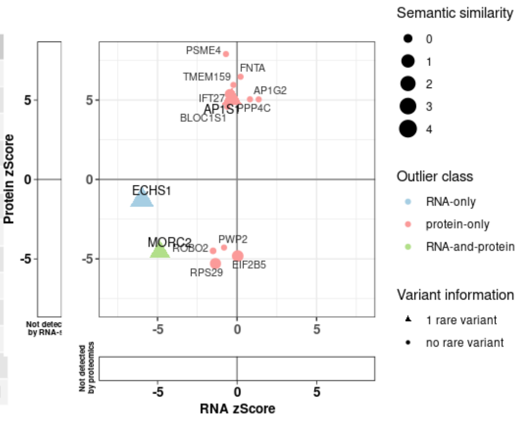


Infant female diagnosed with de-novo heterozygous missense variant (c.[77C>T], p.[Ala26Val]) in *MORC2*. Phenotype of the patient matched the published phenotype of the causal gene (Semantic similarity score = 2.91). MORC2 was a significant RNA-and-protein underexpression outlier (RNA: FC: 0.73; p-value: 3.90*10-6; z-score: -4.86 / Protein: FC: 0.69; p-value: 5.04*10-6; z-score: -4.56).

#### Case 10, Sample ID: OM03592, Causal gene: *MRPL44*

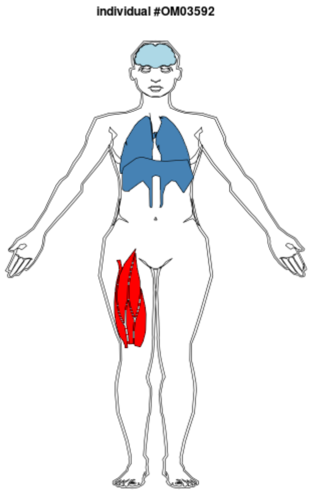


| | HPO_term |
|---|---|
| 1 | Muscular hypotonia |
| 2 | Global developmental delay |
| 3 | Abnormal facial shape |
| 4 | Respiratory distress |
| 5 | Infantile onset |

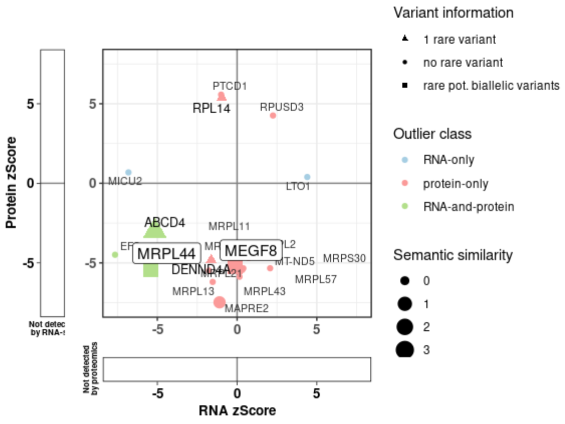


Infant female diagnosed with homozygous nearsplice variant (c.[179+3A>G], p.[?]) in *MRPL44*. MRPL44 was a significant RNA-and-protein underexpression outlier (RNA: FC: 0.74; p-value: 6.68*10-8; z-score: -5.42 / Protein: FC: 0.50; p-value: 6.44*10-8; z-score: -5.41). Splice defect resulting in several aberrant species: truncation and elongation of exon 1 and retention of intron 1. Furthermore reduction of large mitochondrial ribosomal subunit was observed. Although phenotypic presentation was atypical (Semantic similarity score = 1.54), sufficient evidence of pathogenicity allowed to provide the diagnosis and extend phenotypic spectra of the associated disorder.

**Case 11, Sample ID: OM87369, Causal gene: *NDUFB11***

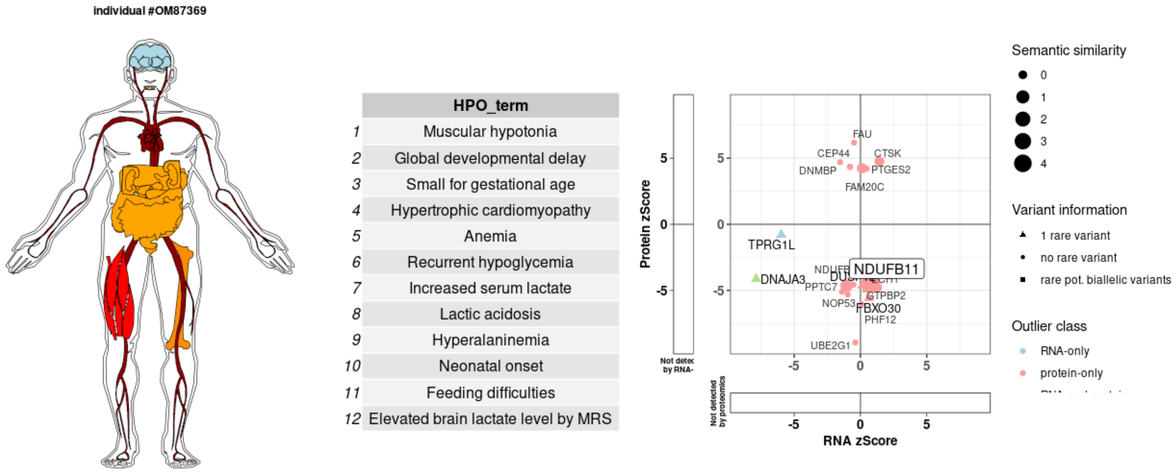


Infant male diagnosed with hemizygous missense variant (c.[440T>C], p.[Met147Thr]) in *NDUFB11*. Phenotype of the patient matched the published phenotype of the causal gene (Semantic similarity score = 3.96). NDUFB11 was a significant protein-only underexpression outlier (Protein: FC: 0.46; p-value: 6.12*10-6; z-score: -4.52). One further significantly reduced protein outlier NDUFB10 (Protein: FC: 0.45; p-value: 3.20*10-6; z-score: -4.66) without potentially biallelic variants as well as a global reduction of respiratory chain complex I was identified.

**Case 12, Sample ID: OM36526, Causal gene: *MRPS25***

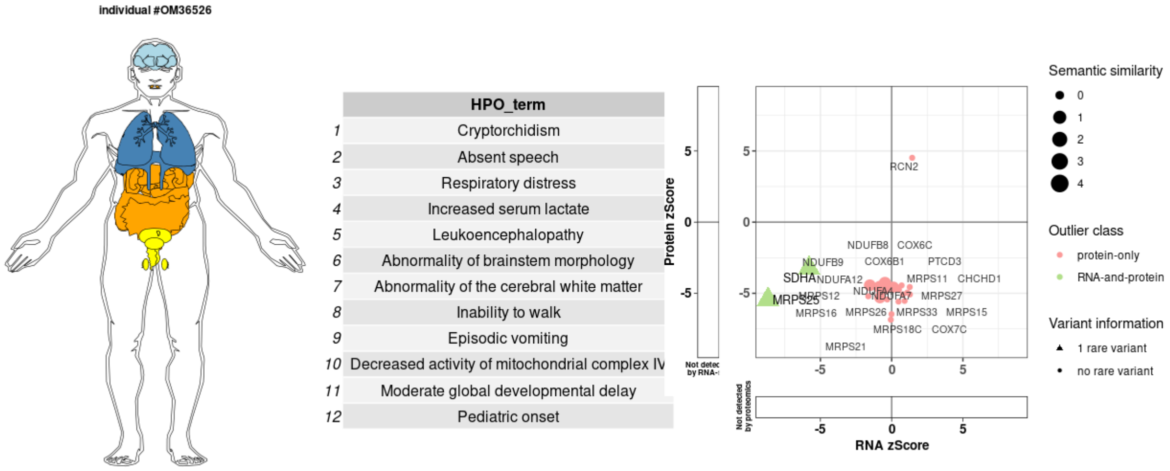


Paediatric male diagnosed with homozygous variant (c.[329+75G>A], p.[Glu111*]) in *MRPS25*. Phenotype of the patient matched the published phenotype of the causal gene (Semantic similarity score = 4.17). Homozygous intronic variants caused exon extension and premature termination, resulting in significant RNA-and-protein underexpression outlier (RNA: FC: 0.43; p-value: 7.68*10-16; z-score: -8.68 / Protein: FC: 0.42; p-value: 5.66*10-8; z-score: -5.43). Furthermore reduction of small mitochondrial ribosomal subunit, as well as respiratory chain complexes, was observed.
